# Human mobility and poverty as key drivers of COVID-19 transmission and control

**DOI:** 10.1101/2020.06.04.20112417

**Authors:** Matan Yechezkel, Amit Weiss, Idan Rejwan, Edan Shahmoon, Shachaf Ben-Gal, Dan Yamin

**Author notes:** To whom correspondence should be sent: Dan Yamin, PhD. These authors contributed equally to this work (Matan Yechezkel, Amit Weiss, Idan Rejwan).

## Abstract

**Background:** Applying heavy nationwide restrictions is a powerful method to curtail COVID-19 transmission but poses a significant humanitarian and economic crisis. Thus, it is essential to improve our understanding of COVID-19 transmission and develop more focused and effective strategies. As human mobility drives transmission, data from cell phone devices can be utilized to achieve these goals.

**Methods:** We analyzed aggregated and anonymized mobility data from the cell-phone devices of>3 million users between February 1, 2020, to May 16, 2020 – in which several movement restrictions were applied and lifted in Israel. We integrated these mobility patterns into age-, risk- and region-structured transmission model. Calibrated to coronavirus incidence in 250 regions covering Israel, we evaluated the efficacy and effectiveness in decreasing mortality of applying localized and temporal lockdowns (stay-at-home order).

**Results:** Poorer regions exhibited lower and slower compliance with the restrictions. Our transmission model further indicated that individuals from poverty areas were associated with high transmission rates. Model projections suggested that, counterintuitively, school closure has an adverse effect and increases COVID-19 mortality in the long run, while interventions focusing on the elderly are the most efficient. We also found that applying localized and temporal lockdowns during regional outbreaks reduce mortality compared to nationwide lockdowns. These trends were consistent across vast ranges of epidemiological parameters, possible seasonal forcing, and even when we assumed that vaccination would be commercially available in 1-3 years.

**Conclusions:** More resources should be devoted to helping impoverished regions. Utilizing cellphone data despite being anonymized and aggregated can help policymakers worldwide identify hotspots and apply designated strategies against future COVID-19 outbreaks.

## Background

Severe acute respiratory syndrome coronavirus 2 (SARS-CoV-2) was identified in Wuhan, China, in December 2019. It has since developed into a pandemic wave affecting over 200 countries, causing over 6.9 million cases and claiming over 390 thousand lives, as of June 8, 2020 [1]. The rapid growth of the SARS-CoV-2 pandemic led to unprecedented control measures on a global scale. Travel bans, restrictions on mobility of varying degrees, and nationwide lockdowns have emerged sharply in over 200 countries [2]. In Israel, since March 9, 2020, travelers from any country are being denied entry unless they can prove their ability to remain under home isolation for 14 days. From March 16 onward, daycare and schools were shut, and work was limited to less than a third of the capacity. On March 26, inessential travel was limited to 100 meters away from home, and three lockdowns were applied in most regions in Israel to prevent crowding due to holiday celebrations [3].

These massive measures have led to a sharp decline in transmission but pose a significant humanitarian and economic crisis [4–7]. Recent estimates have suggested that 1.5-3 month lockdowns will lead to an enormous economic loss, with high variability across countries ranging between 1.7-13.1% decline in the gross domestic product[4]. Restrictions to mitigate the outbreak also led to various types of psychological distress, including anxiety, helplessness, and depression [5–7]. Furthermore, social isolation is a primary public health concern in the elderly, as it also amplifies the burden of neurocognitive, mental, cardiovascular, and autoimmune problems [7]. Thus, given that pandemics rarely affect all people in a uniform manner [8], it is essential to improve our understanding of the COVID-19 transmission dynamics to customize control efforts.

As human mobility is an intrinsic property of human behavior, it serves as a key component of the transmission of respiratory infections, including COVID-19 [9–13]. The four billion mobile phones in use worldwide are ubiquitous sensors of individuals’ locations and can be utilized not only to track mobility patterns, but also to understand compliance with ongoing restrictions [12]. The importance of human mobility is further intensified by the 2.2-11.5 days of incubation, and the observation that as many as 95% of cases are unreported [14]. Thus, utilizing real-time data on human mobility is instrumental for early detection and prompt isolation of COVID-19 infection.

A variety of factors besides human mobility affect the risk of infection and manifestations, including demographics, education, underlying conditions, and epidemiological characteristics [15]. The high variance in the severity of the disease for different age groups suggests that age-based strategies might be useful in reducing mortality [16]. Age-stratified modeling studies show that interventions such as school closure can help delay the outbreak peak [11]. However, this will not necessarily result in a reduction in the total number of deaths, particularly in light of the estimated time for vaccine availability being >1 year [17]. In addition to age, individuals with comorbidities are 2.8-21.4 times more likely to become hospitalized following COVID-19 infection [18]. Another factor may be socioeconomic status. Impoverished populations often live in denser regions and have reduced access to health services, thereby being most vulnerable during a crisis [8]. The considerably high rate of household transmission for respiratory infections [19] may also suggest a higher risk for larger families, regardless of lockdowns.

We analyzed a large-scale data of location records from mobile phones to explore the spatiotemporal effect of human mobility and population behavior on transmission. We integrated these mobility data into regional age- and risk-structured transmission model and used our model to identify efficient and effective strategies for reducing COVID-19 mortality. Our methodology can help policymakers worldwide utilize aggregate and anonymized cellphone data to develop designated strategies against future outbreaks.

## Methods

### Human mobility

Our data include mobility records based on cellular data of >3 million users from one of the largest telecommunication companies in Israel. With the exception of children <10 years of age, the users are well representative of Israel demographically, ethnically, and socioeconomically. In accordance with the General Data Protection Regulation (GDPR), the data include aggregated and anonymized information. The data specifies movement patterns within and between 2,630 zones covering Israel, on an hourly basis, from February 1, 2020, until May 16, 2020. To ensure privacy, if less than 50 individuals were identified in the zone in a given hour, the number of reported individuals was set to zero.

We determined the location of individuals based on the triangulation of cell towers, which was found to be accurate to 300 meters in most cases but varied by up to 1 km in less populated areas. To prevent signal noise and identify stay points, we tracked only locations where users stayed for at least 15 minutes within a distance threshold of 1.5 km. We defined users as residents of a zone based on the location at which they had the highest number of signals on most nights during February 2020. We define a mobility index (MI) as the daily proportion of individuals who traveled >1.5 km away from their home. To calculate the MI for each zone, we counted the daily number of individuals in each group that showed a signal away from their home location.

Next, we integrated data from the Central Bureau of Statistics (CBS) that specifies several socioeconomic characteristics, including population size, household size, age distribution, socioeconomic score, and dominant religion, for each zone. Each zone includes ∼3,500 residents. For each zone, we scaled the number of resident users of the telecommunication company to match the actual number of residents in the zone, as reported by the Israeli CBS. The CBS specifies for each zone a socioeconomic cluster from 1 to 10. Based on these clusters, we defined three SES groups that were nearly equal in size: low (clusters 1-3), middle (clusters 4-7), and high (clusters 8-10). We aggregated the MI according to SES to test the mobility trends on a national level (Fig. 1A). To evaluate the travel patterns based on an individual’s SES (Fig. 1B and 1C), we counted the mean daily number of travels between the 2,630 zones, including for those individuals who stayed in their origin zone. Grouping by SES and scaling the daily number of travels to one for each zone, we created an origin-destination travel probability matrix.

**Fig. 1.**
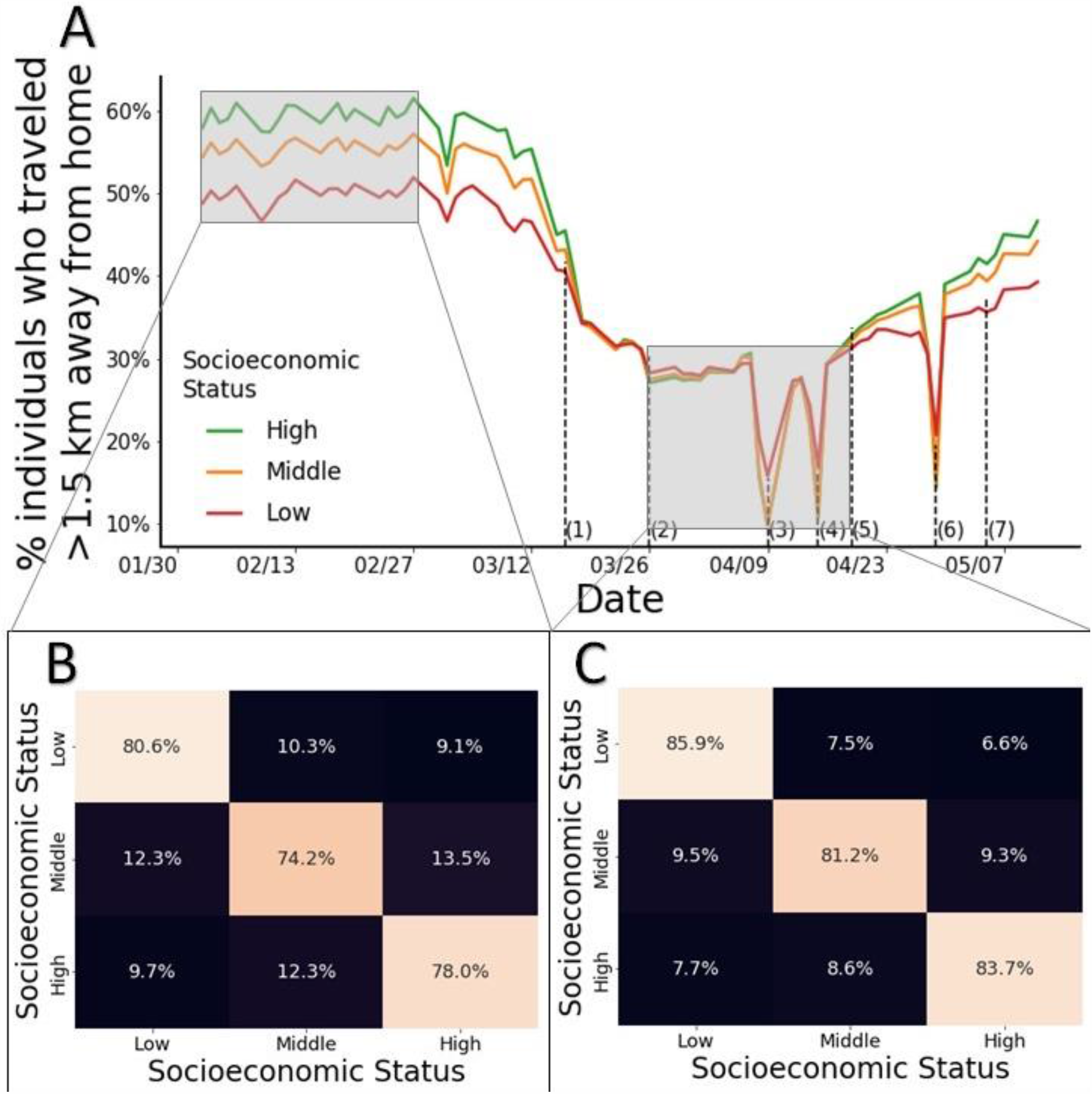
Mobility patterns with and without restrictions. (A) Percentage of individuals who traveled >1.5km, stratified by socioeconomic groups, during routine and when mobility restrictions were applied and lifted: (1) closing schools and stores and limiting workplaces to 30% activity; (2) limiting nonessential travels to 100 meters away from home; (3) and (4) national daily lockdowns due to Passover; (5) opening stores; (6) lockdown due to Independence Day; (7) lifting the 100 meter limit for nonessential travels. (B) and (C) Travel patterns based on individuals’ SES during February 2-29 (B) and March 26-April 18 (C).

To analyze the relationship among poverty, mobility, and transmission (Fig. 2), we divided the data into three periods: 13 Feb-26 Mar, 27 Mar-19 Apr, and 20 Apr-15 May, corresponding to 1) the early phase before restrictions started, 2) the time from restrictions until they were first lifted, and 3) after the restrictions were lifted. For each period, we ranked municipalities with a population of >10,000 residents based on the number of new cases per person observed in each period. For improved clarity of Fig. 2, we present the 50 most prevalent municipalities. We calculated for each city the number of newly reported cases, the SES, and the distribution of travels to the other 49 municipalities.

**Fig. 2.**
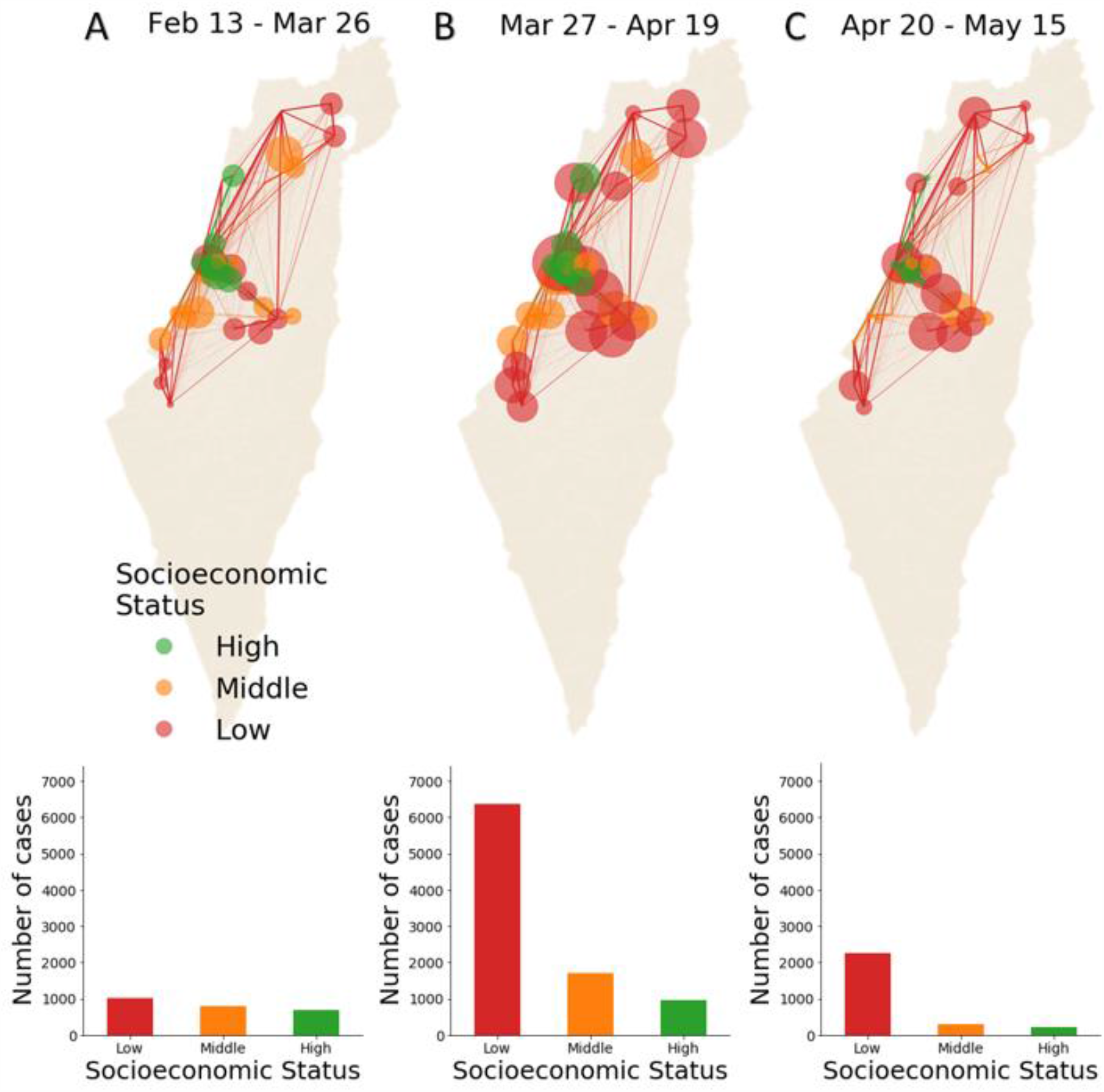
Association between mobility and poverty in COVID-19 transmission. Spatiotemporal transmission by socioeconomic status. We present the 50 municipalities with the highest incidence. Each circle represents one municipality. The radius (presented on a logarithmic scale for clarity) reflects the total number of new cases reported during the corresponding period. The colors reflect socioeconomic status. The lines between the municipalities represent the traffic of each municipality, wherein the line thickness represents the relative traffic intensity and the color matches the color of the SES of origin. We present below each map the number of reported cases among different SEGs for three periods corresponding to (A) the early phase before restrictions started, (B) from the time of restrictions and until the restrictions were lifted, and (C) after restrictions were lifted.

### Transmission model

We developed a dynamic model for age-, risk- and region-stratified SARS-CoV-2 infection progression and transmission in Israel. Our model is a modified susceptible exposed infected recovered (SEIR) compartmental framework [20], whereby the population is stratified into health-related compartments, and transitions between the compartments change over time (Fig. 3A). To model age-dependent transmission, we stratified the population into age groups: 0-4 years, 5-9 years, 10-19 years, 20-29 years, 30-39 years, 40-49 years, 50-59 years, 60-69 years and ≥70 years. We distinguished high-risk and low-risk individuals in each age group based on the ACIP case definition [21, 22]. We also distinguished the 250 regions covering Israel in the model.

**Fig. 3.**
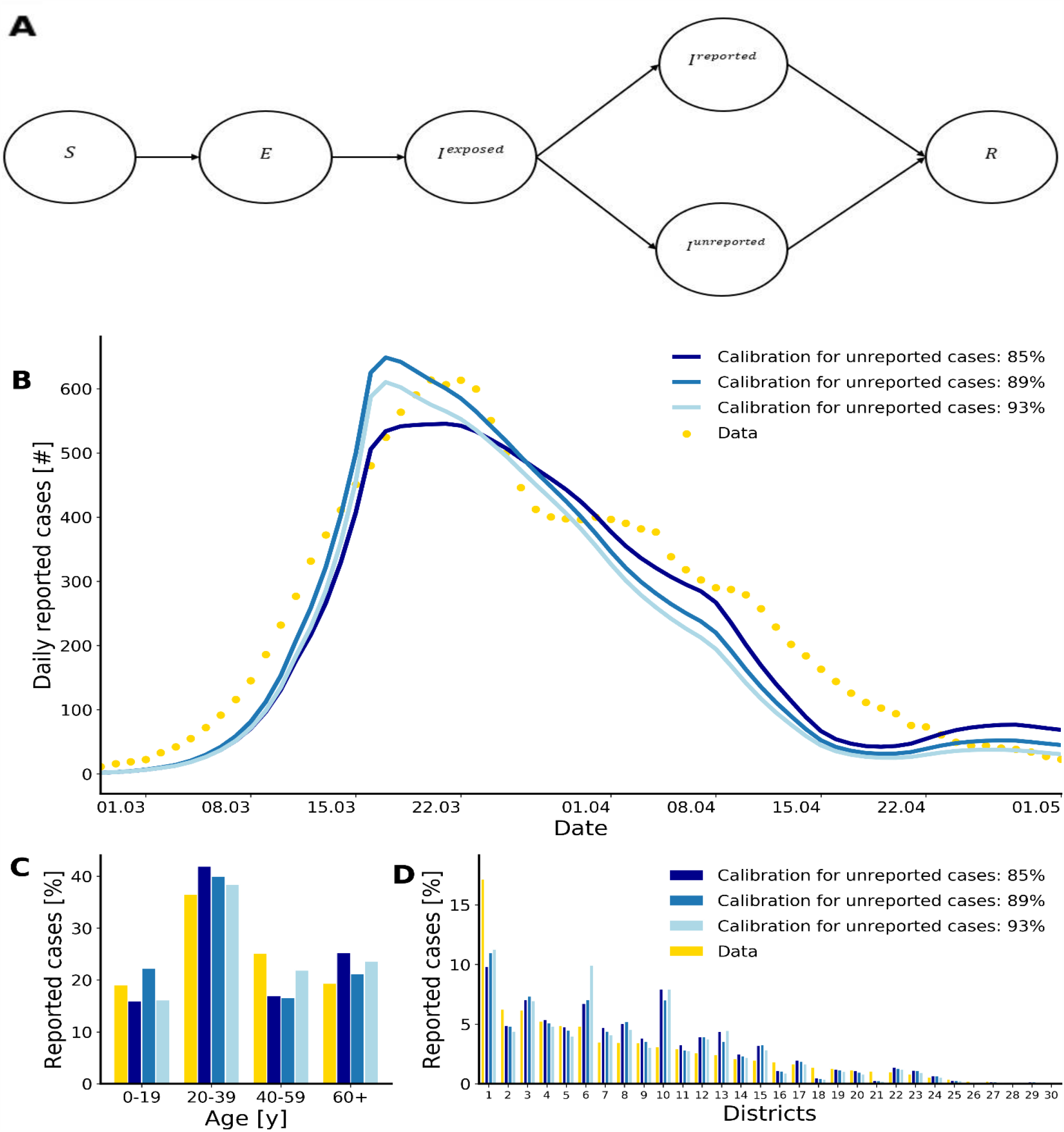
Structure and fit of the transmission model. (A) Compartmental diagram of the transmission model. Susceptible individuals *S* transition to the exposed compartment with a force of infection λ, where they are infected but not yet infectious, until moving to an early infectious compartment at rate *σ*, in which they do not show symptoms but may transmit. Infected individuals in the early stage move to a reported *I*^*Reported*^ or unreported *I*^*Unreported*^ infectious period, in which they may have a mild or an asymptomatic infection until death or complete recovery. For clarity of depiction, age, risk, and region stratifications are not displayed. (B) Time series of reported daily COVID-19 cases and model fit countrywide. (C) Data and model fit to the age distribution among COVID-19 infections. (D) Data and model fit to the 30 subdistricts covering Israel.

The mean incubation period of SARS-CoV-2 is 6.4 days (95% CI, 5.6 to 7.7 days) [23, 24], but early evidence shows that viral shedding occurs during a presymptomatic stage [25, 26]. Thus, we considered an exposure period *E* and an early infectious period *I*^*exposed*^. Underreporting arises from asymptomatic cases or mild cases in individuals who do not seek care. Thus, following the early infectious phase, individuals in the model transition either to an infectious and reported compartment *I*^*reported*^ or to an infectious and unreported compartment *I*^*unreported*^ [27, 28].

Multiple infections with SARS-CoV-2 are not yet fully understood. A recent study indicated that there is protective immunity following infection [29]. This result is consistent with a previous study indicating that for SARS-CoV-1, memory T cells persist for up to 11 years [30]. In addition, similar to other respiratory infections, it is likely that if reinfection occurs, it is less severe and less transmissive [31]. Thus, we assumed that upon recovery, individuals are fully protected, which is consistent with other SARS-CoV-2 transmission models [32] (Additional file 1: Supplementary information). Altogether, our model includes 5 ∗ 9 ∗ 2 ∗ 250 *=* 22,500 compartments (*health* − *compartments* ∗ *age* − *groups* ∗ *risk* − *groups* ∗ *regions)*.

### Force of infection and seasonality

The rate at which individuals transmit depends on (i) contact mixing patterns between the infected individual and his or her contact, (ii) age-specific susceptibility to infection, (iii) region-based behavioral susceptibility, and (iv) potential seasonal forcing.

Age-specific contact rates were parameterized using data from an extensive survey of daily contacts [33] and data from CBS regarding the household size in each region. In addition, we utilized the aggregate mobility data regarding movement patterns within and between 250 regions as observed in the data during routine and following restrictions (Additional file 1: Supplementary information). We specifically distinguished the contact patterns of infected individuals for different locations, namely, at home, at work and during leisure, such that the number of contacts was based on the extensive survey [33] and the household size, whereas the mixing patterns were based on the locations of the individuals as analyzed using the mobile data. These contact data reveal frequent mixing between similar age-groups, moderate mixing between children and people their parents’ age, and infrequent mixing among other groups. The data based on mobility reveal more frequent mixing between individuals of similar SES, at similar geographical distances, and with cultural similarities (Additional file 1: Supplementary information).

We distinguished between in-home and out-of-home transmission. We evaluated the in-home transmission is independent of age, and based on a previous retrospective studies, that suggested a value of 0.16 [19]. The age-specific susceptibility rate for out-of-home individuals *β*_*j*_ was parameterized by calibrating our model with daily COVID-19 records.

To account for behavioral susceptibility, we explicitly considered in our model a parameter reflecting the order to maintain physical distancing, *κ*_*p*_. The high regional variations in susceptibility were parameterized based on fertility rates and socioeconomic characteristics. Specifically, we computed for each region the relative change in mobility compared to routine. Our analysis indicated that for regions of low SES, the change was lower, which was reflected in our model by higher susceptibility (Additional file 1: Supplementary information). The use of regional fertility and relative change in mobility allowed us to refrain from calibrating the model to an excessive number of unknown parameters and avoid overfitting.

Seasonal patterns have been observed in common circulating human coronaviruses (HCoVs), mostly causing infections in humans between December and May in the Northern Hemisphere [34]. The two HCoVs 229 E and OC43 show distinct winter seasonality. In addition, many coronaviruses in animals exhibit a distinct seasonal pattern of incidence in their natural hosts [35]. There is growing evidence that SARS-CoV-2 is also seasonal, with the optimal setting for transmission in Israel occurring during winter [36]. Thus, we considered in our base-case seasonal forcing by including general seasonal variation in the susceptibility rate of the model as

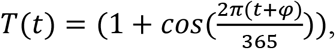

in which *φ* is the seasonal offset. This formulation was previously shown to capture the seasonal variations in several respiratory infections, including RSV and influenza [31, 37]. We incorporated the possible values of *φ* to reflect peaks from December through February (Additional file 1: Supplementary information).

### Model calibration

To empirically estimate unknown epidemiological parameters (Additional file 1: Table S5), we calibrated our model to daily age-stratified cases of COVID-19 confirmed by PCR tests in 30 subdistricts covering Israel. The calibration was conducted on a 30-subdistrict level rather than in the 250 regions to ensure that there were sufficient time series data points in each location for each age-group. The data were reported by the Israeli Ministry of Health between February and May and include daily information for the patients, including age, residential zone, underlying conditions, and clinical outcomes, including hospitalizations and death.

Due to the uncertainty regarding the proportion of unreported cases, we calibrated our model to different scenarios. Specifically, underreporting is affected by testing policy and testing capabilities for each country, as well as individuals’ tendency to seek care once clinical symptoms appear. In addition, underreporting is affected by the severity of the infection, which is associated with age [18]. Thus, we chose different estimates for the proportion of underreporting, ranging from 5.5-14 unreported cases for a single reported case. These estimates are based on observations from screenings conducted in unpublished data from Israel and are consistent with data from Denmark, Czechia, Netherlands; Santa Clara, California [14, 18, 38] (Additional file 1: Table S1). Due to the uncertainty related to positive predictive values of serological screenings, we also tested a scenario of two unreported cases for a single reported case to confirm the robustness of our findings.

To account for the age variation, we considered the detailed serological data from Santa Clara [14]. We also calibrated our model with scenarios assuming different phases of seasonal peaking between December 21 and February 21, as well as scenarios with no seasonality. The final transmission model included five parameters without constraints imposed from previous data: reduced susceptibility due to physical distancing *κ*_*p*_ and susceptibility rate based on age groups *j*: 0-19, 20-39, 40-59, and >60 (Additional file 1: Supplementary information).

### Model simulations

We evaluated the effectiveness of temporal lockdown strategies in reducing mortality by simulating the model for one year and three years or until disease elimination. Each strategy considered includes a threshold for activation of a lockdown, and the groups considered for lockdown were as follows: 1) the entire population in the region, 2) daycare- and school-age children between 0-19 years of age (children), 3) high-risk groups and individuals >65 years of age (elderly). Specifically, to model the lockdown strategies, we defined an indicator for each region as the weekly number of new-reported cases per 10,000 people. Each week, we examined whether the indicator exceeds a certain threshold for each region. If so, a lockdown was activated for the following week. This process was continued for 1-3 years.

We simulated the lockdowns in our model based on the mobility patterns we observed between March 26 and April 16 during which a stay-home orders were applied. In this period, school and daycare centers were closed, and for non-essential workplace only 10% of employees from private and public sectors were allowed to work. Individuals were required to stay in a radius of 100 meters from their home except for grocery and health-related shopping.

We projected the number of individuals who will die under each strategy by utilized available detailed information from the Israeli Ministry of Health (Additional file 1: Table S2). Specifically, we calculated for each age- and risk-group the proportion of individuals who died out of the reported cases. We multiplied these proportions with the daily model projections of newly reported cases and summed this product to calculate the total projected number of deaths. We also accounted for the uncertainty regarding the estimated probabilities. We define the efficiency of a lockdown strategy as the total number of deaths averted per total lockdown days. The number of deaths averted is calculated as the projected number of deaths with no lockdowns minus the number of deaths projected when the considered strategy is applied.

## Results

### Human mobility and poverty

We utilized aggregated and anonymized information about mobility based on cellular data. The data specifies movement patterns of >3 million users within and between 2,630 zones covering Israel, on an hourly basis, from February 1, 2020, to May 16, 2020. This period corresponds to the period from a month before the COVID-19 outbreak began in Israel until 16,600 cases were reported. Each zone includes ∼3500 residents with available information regarding several socioeconomic characteristics, including household size, age distribution, mean socioeconomic score, and religion.

During the aforementioned period, the government applied and lifted several movement restrictions. We define a mobility index (MI) as the daily proportion of individuals who traveled >1.5 km away from their home. While a sharp decline has been observed in the overall population following restrictions, the decline varied considerably among individuals of different socioeconomic statuses (SESs). Specifically, during routine days, the low-SES population had the lowest MI. Shortly after the restrictions started, this trend changed, and populations of all SESs had similar MIs, while during the lockdowns, the high-SES population had the lowest MI (Fig. 1A).

Before the COVID-19 outbreak, the population was highly clustered such that people of a specific SES typically traveled to zones where the residents matched their SES and were therefore more likely to meet with each other (Fig. 1B; Additional file 1: Figs. S1 and S2). Likewise, people of similar demographic groups, such as those with the same religious affiliations, typically traveled to zones where the residents matched their group. These trends further intensified following the restrictions (Fig. 1C). Notably, the clustering was not attributable to only the geographical distance, as many high-SES zones are geographically close to the low-SES zone.

### Human mobility and poverty explain transmission

To explore the spatiotemporal effect of human mobility and poverty on transmission, we calculated the number of new cases and the amount of travel between zones observed during three periods: February 13-March 26, March 27-April 20, and April 20-May 20 (Fig. 2). These periods correspond to 1) the early phase before restrictions started, 2) between the time of restrictions and until the restrictions were lifted, and 3) after restrictions were lifted. Our analysis indicated that during the first period, the infection was evenly distributed among different SESs. During the second period, 71% of the cases were residents of zones with a low SES, particularly religious orthodox Jews. During the third period, 81% of the cases were residents of low SES, mainly residents of zones of Israeli Arabs and orthodox Jewish people. We also identified a high correlation ranging from 79.2-82.8% (p value<0.001) with a lag of 12-14 days between the MI and the disease growth factor, i.e., the number of new cases daily per active case (Additional file 1: Fig. S3). This lag includes the incubation period, the time from symptom onset until a test is conducted, and the time until the test results arrive.

We integrated the daily mobility data into an age-, region-, and risk-stratified model for SARS-CoV-2 transmission. Model parameters were calibrated to the number of new cases daily in 30 subdistricts covering Israel. With only five free parameters, the model recapitulated SARS-CoV-2 trends (Fig. 3). For example, the calibrated model showed that the national SARS-CoV-2 infections peaked during March 17-25 (Fig. 3B) and yielded age and regional distributions of SARS-CoV-2 consistent with the data (Fig. 3C and D). Our calibration further indicated that a model ignoring mobility poorly captured the spatiotemporal dynamics and provided overestimation of disease transmission (Additional file 1: Table S5). We also found that a model that accounted for seasonal forcing yielded a higher, but not significant (p value<0.35), likelihood than a model that did not account for seasonal forcing (Additional file 1: Table S5).

### Focused lockdowns reduce mortality

As transmission varied considerably among regions, we projected the number of total deaths for 1-3 years under local and temporal lockdown strategies. Specifically, we simulated three strategies triggered by a threshold of daily COVID-19 incidence in each of the 250 regions. We evaluated the efficiency of the lockdown strategies, defined as the number of deaths averted per lockdown day (Fig. 4). We found that the local strategy of targeting the elderly was substantially more efficient than nationwide strategy. For example, assuming the proportion of unreported cases is 85% and a lockdown threshold of 5/10,000 (cases/individuals), a strategy targeting the elderly is 4.3-5.5 times more efficient than a global strategy (Fig. 4C and D).

**Fig. 4.**
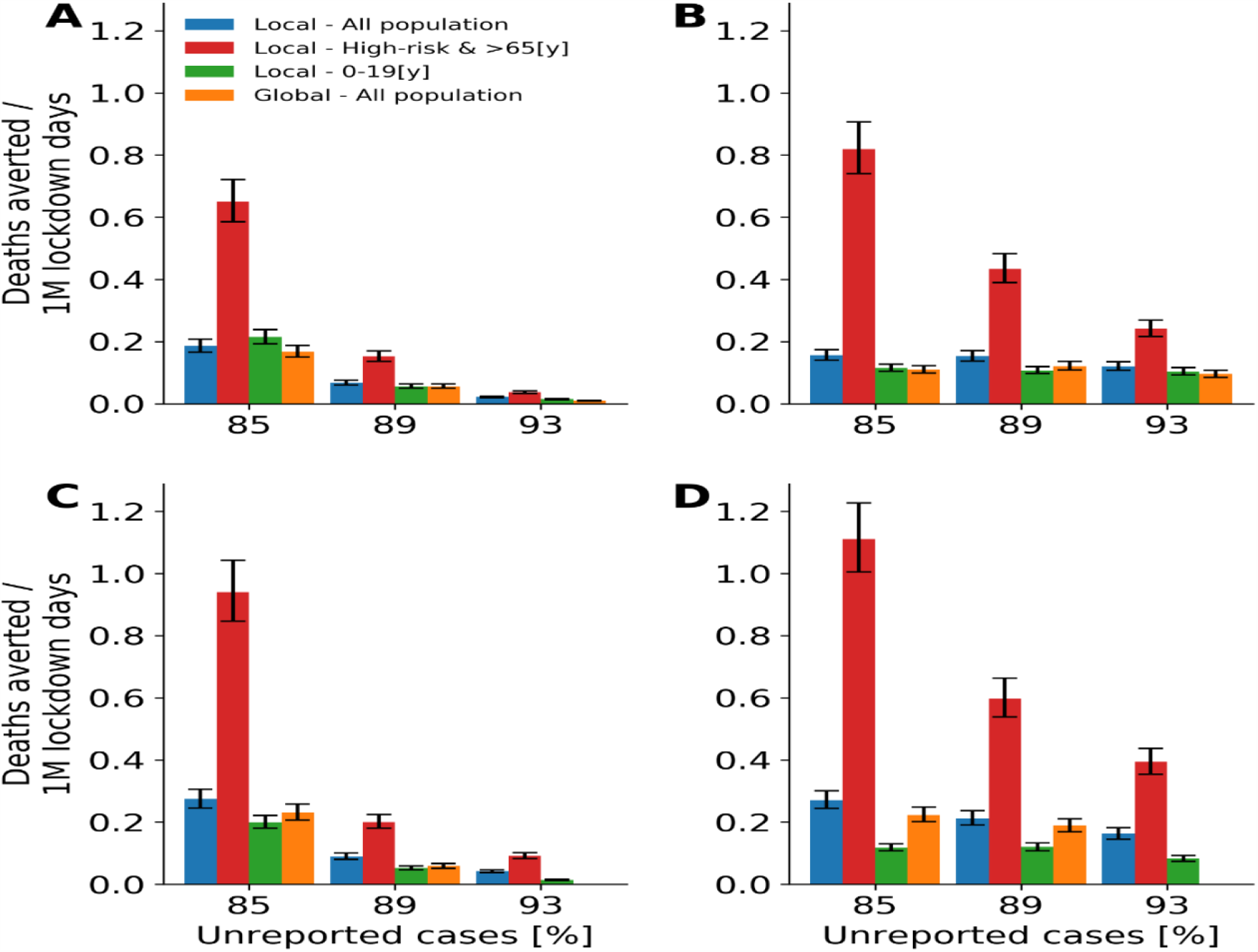
Efficiency of lockdown strategies. Median and interquartile values of the projected number of deaths averted per 1 million lockdown days due to the implementation of lockdown strategies (A, C) after one year and (B, D) after three years. (A, B) The thresholds for lockdowns in a local region are 1/10,000 [cases/individuals] and (C, D) 5/10,000 [cases/individuals].

We evaluated the effectiveness of each strategy in reducing mortality (Fig. 5). We found that a strategy locally targeting the elderly yielded a lower number of deaths than a strategy targeting children. For example, assuming the proportion of unreported cases is 85% and a lockdown threshold of 5/10,000 (cases/individuals), a strategy targeting the high-risk group resulted in 4,500-4,900 deaths while on targeting children resulted in 7,900-10,500 deaths after one year (Fig. 5A and C). In addition, for lockdown thresholds exceeded 5/10,000, which aligns with the current practice in Israel, a strategy locally targeting the elderly either is projected to be the most effective or is comparable to the most effective strategies. Although comparable on the effectiveness, such a policy includes 2.2-5.5 times fewer individuals under lockdowns (Fig. 5C and D). These trends were consistent across vast ranges of epidemiological parameters, different plausible ranges of threshold values, and different considerations of seasonal forcing.

**Fig. 5.**
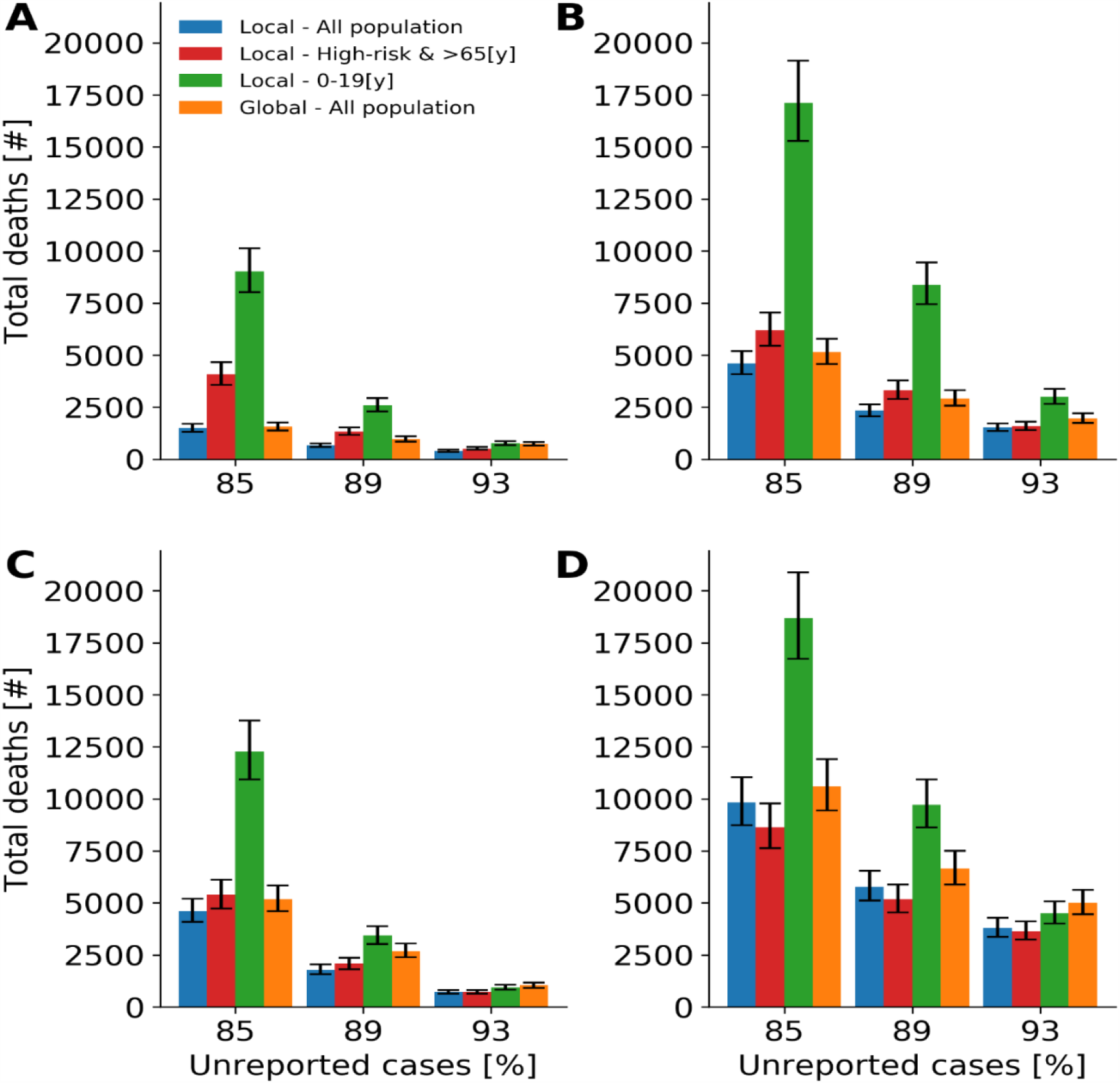
Effectiveness of lockdown strategies. Median and interquartile values of the projected number of deaths after implementation of strategies (A, C) after one year and (B, D) after three years. (A, B) The thresholds for lockdowns in a local region are 1/10,000 [cases/individuals] and (C, D) 5/10000 [cases/individuals].

## Discussion

Our key findings suggest that COVID-19 infection does not spread uniformly in the population, and thus, intervention strategies should focus primarily on protecting elderly and individuals with underlying conditions in regions of outbreaks. Such a strategy can reduce mortality while enabling daily routine for a vast majority of the population.

Our work demonstrates that to understand the spatiotemporal dynamics of transmission, models must account for mobility as well as behavioral aspects that are associated with sociodemographic and socioeconomic factors. In particular, we found that SARS-CoV-2 is more likely to spread in more impoverished regions and is affected by human mobility. The intensive interactions likely led to higher transmission in developed countries than in developing countries. However, our model suggested that people of low SES are at higher risk due to poorer compliance and larger household size. Thus, to contain the COVID-19 outbreak more resources should be devoted to helping improvised regions.

Our analyses indicate that localized lockdowns with incidence thresholds as low as five reported cases in 10,000 individuals are essential to decrease mortality. This finding underscores the importance of maintaining a high level of testing [39], particularly in regions with elevated risk of transmission. However, with such a strategy, at least 2500 total years of lockdowns (equivalent to a one-day lockdown of 912,500 individuals) are required to prevent a single death. Considering that one day of lockdown is equivalent to a quality of life value that is ∼0.85 times that in a routine day [40], even local lockdowns should be prudently considered from a health economic perspective. Thus, future modeling studies should also include localized and temporal massive screening efforts, which result in more focused quarantines and isolations than massive control measures.

As in any modeling study, we made several assumptions. Nationwide and local lockdowns are powerful, yet heavy, control measures. Thus, the local strategies tested in our model should be applied only if containment cannot be achieved via less drastic measures to the economy such as the use of contact tracing to break the chains of infection, requiring the use face masks and educating to maintain physical distancing. We denote that these measures were applied in Israel and were taken into consideration in our model indirectly by our calibration process. Thus, our model suggests the disease cannot be contained by these measures in the extent they were implemented.

We assumed in our model that there is a long-lasting protective immunity following infection which is consistent with previous human coronavirus types [29, 30, 32]. However, a recent study suggested that people are unlikely to produce long-lasting protective antibodies against this virus [41]. If, indeed, a rapid waning is possible, this highlights the importance to protect the elderly in regions of high outbreaks.

Our local lockdowns correspond to regions with a population of ∼36,000 people. A smaller lockdown may be more efficient but could not be tested by our model. Additionally, with the growing evidence of a disproportionate risk from COVID-19 to the elderly [18, 42], focused control measures are likely to be conducted in retirement homes and facilities with populated communities at high risk, which we did not explicitly account for in our model [43]. Although the transmission dynamics are unlikely to change with such focused interventions, the overall mortality is expected to be lower than what we have found.

While there is a debate in the literature regarding the extent of infectiousness and transmissibility in children [44], our results highlighted a not less important question: to whom do children transmit? Our findings reveal that children are less likely to transmit to populations at risk, and thus, a differential lockdown strategy that targets children may be even harmful.

## Conclusion

We showed that using aggregated and anonymized human mobility data from cellular phones under the General Data Protection Regulation (GDPR) guidelines is a powerful tool to improve the understanding of transmission dynamics and to evaluate the effectiveness of control measures. Our transmission model predicted that rather than nationwide lockdowns, applying temporal and localized lockdowns that focus on elderly can substantially reduce mortality. Such focused measures will enable a vast majority of the population to maintain a daily routine.

## Data Availability

Data is available upon request

## List of abbreviations

MI: Mobility Index
GDPR: General Data Protection Regulation
SES: Socioeconomic Status
CBS: Central Bureau of Statistics

## Declarations

### Ethic approval

We hereby declare that the IRB chair of Tel Aviv University, Prof. Meir Lahav, determined on March 24, 2020, that there is no need for an IRB approval for this study. We also received consent from the data provider to use the aggregated and anonymized human mobility data in the way it is used in our study.

### Consent for publication

Not applicable.

### Data and materials availability

The medical data that support the findings of this study are publicly available by the Israeli Ministry of Health, https://data.gov.il/dataset/covid-19. The aggregated and anonymized human mobility data which were used under the license for the current study are not publicly available. Data, however, available from the authors upon reasonable request.

### Code availability

The code is available on GitHub.

### Author contributions

DY, MY, AW, IR contributed to the study design, analysis, and interpretation of the results. ES and SB contributed in providing and interpreting the raw data. DY wrote the first draft of the manuscript. All authors contributed to further versions of the manuscript. All authors have read and approved the manuscript.

## Acknowledgments

The authors would like to thank Prof. Irad Ben-Gal, Yariv Waits, Moris Suissa, Dana Pessach, Dganit Meron for their valuable insights on analysis.

## 1. Model

### 1.1. The model

We developed a dynamic model for age-, risk- and regions-stratified SARS-Cov-2 infection progression and transmission in Israel. Our model is a modified Susceptible-Exposed-Infected-Recovered (SEIR) compartmental framework [1], whereby the population is stratified into health-related compartments, and transitions between the compartments occurs over time (Main text, Fig. 3). To model age-dependent transmission, we stratified the population into nine age groups: 0–4 years, 5-9 years, 10-19 years, 20-29 years, 30-39 years, 40-49 years, 50-59 years, 60-69 years and ≥70 years. [2–4]. We distinguished between high-risk and low-risk individuals for each age group based on the ACIP case definition [5,6]. We also distinguish in the model between 250 regions covering Israel.

Multiple infections with SARS-Cov-2 is yet fully understood. A recent study indicated that there is a protective immunity following infection in humans [7] and animals [8]. This result is in-line with a previous study indicating that for SARS-Cov-1, Memory T cells persist for up to 11 years [9]. In addition, similarly to other respiratory infections, it is likely that if re-infection occurs, it is less severe and less transmissive [10]. Thus, we assumed that upon recovery individuals are fully protected for the entire season wich consistent with other SARS-COV-2[11,12].

The mean incubation period of SARS-Cov-2 is 6.4 days (95% CI, 5.6 to 7.7 days) [13,14], but first evidence shows viral shedding occurs during a pre-symptomatic stage [15,16]. Thus, we considered an exposed period *E*, and an early infectious period *I*^*exposed*^. Underreporting arises from asymptomatic cases or mild cases of individuals that do not seek care [17–20]. Thus, following the early infectious phase, individuals in the model transition either to an infectious and reported compartment *I*^*reported*^, or to infectious and unreported compartment *I*^*unreported*^.

To enable in our model for a subset of the population to go for intervention (e.g., 30% of the individuals from specific regions, age groups or risk-group to go under lockdown during a selected time period), we also specifically distinguish between those who undergo and those who did not undergo an intervention.

Accordingly, we stratified the population into six health-related compartments: susceptible *S*_*j,k,r,q*_(*t*), exposed but not yet infectious *E*_*j,k,r,q*_(*t*), infectious at early stage 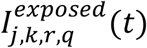, reported infectious 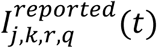, unreported infectious 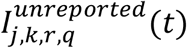 and recovered *R*_*j,k,r,q*_(*t*), such that at any given time t (in days) the population is fixed and scaled to one. Namely,

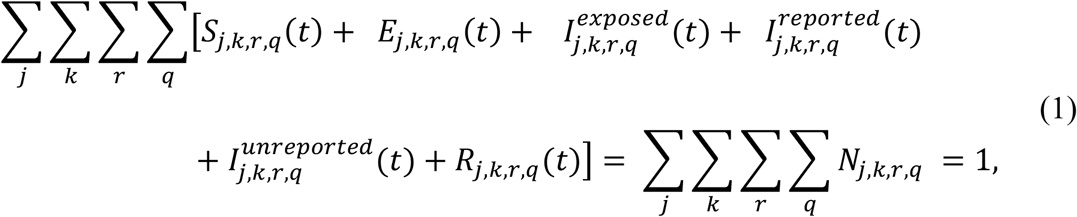

where the index *j* ∈ {0 − 4*y*, 5 − 10*y*, …, > 70*y*} represents the age-group of each individual, index *k* ∈ {1,2, …, 250} specifies the home region of each individual, index *r* ∈ {*L, H*} specifies the risk-group of each individual (i.e. High-risk, or low-risk) and index *q* ∈ {*intervention, non* − *intervention*} represent the intervention-group of each individual.

### 1.2. Model transitioning

Susceptible individuals *S*_*j,k,r,q*_(0), transition to the exposed compartment *E*_*j,k,r,q*_(*t*), with force of infection *λ*_*j,k,q*_(*t*), depending on their age-group *j* home region-group *k* and their intervention-group *q*. At this compartment individuals are infected but not yet infectious until they move at rate *σ* to an infectious compartment 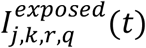, where they are at the early stage of the infectious period. Infected individuals at early stage of their infectious period, then move at rate *δ* to the late infectious period, where they can become to a unreported case (having non to mild symptoms) with probability *f*_*j,r*_ which results in transition to 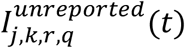. With probability of (1 − *f*_*j,r*_) they can become to a reported case (having moderate to severe symptoms), which results in transition to 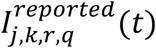. After infectious period, individuals’ transition into the recovered compartment at rate *γ, R*_*j,k,r,q*_(*t*),. (See Section, 2.3 Epidemiological parameters). We also consider a function of the initial spreaders with time *ε_j,k,r_*(*t*), that reflects the individuals exposed to the virus the entered Israel from overseas between February 21 2020 - and March 9, 2020. Thus, the transmission model is composed of the following system of difference equations:

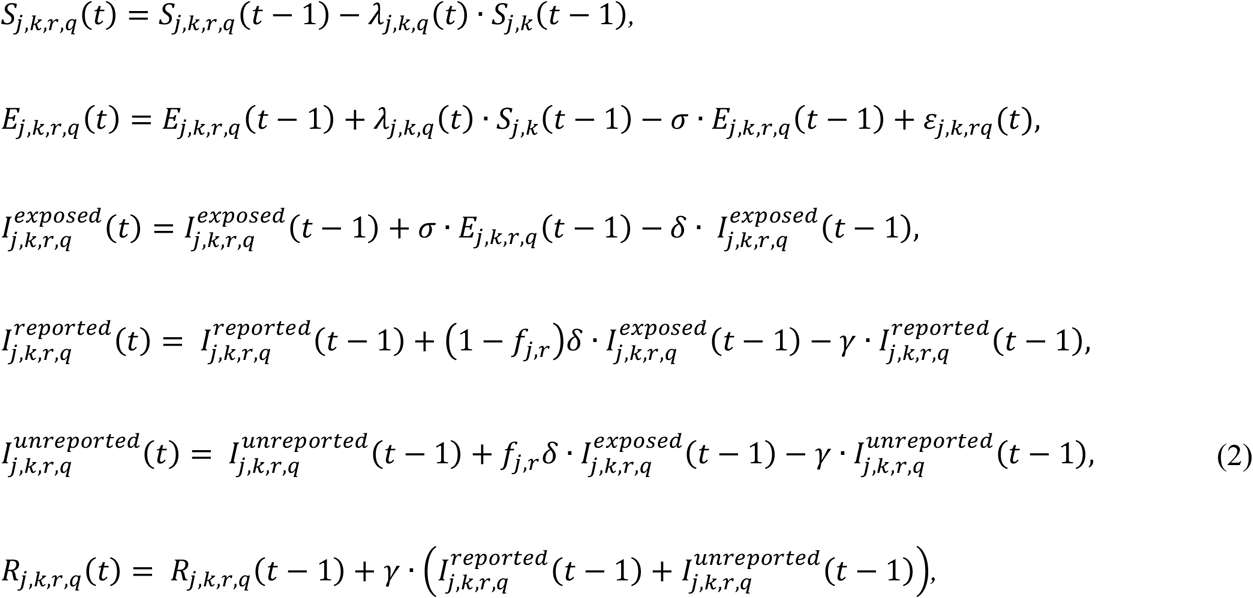

with initial conditions:

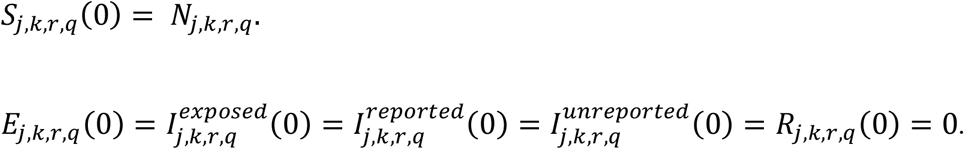

### 1.3. Force of infection

The rate at which individuals transmit SARS-Cov-2 at time t is *λ*_*j,r,q*_(*t*). This rate depends on the combination of (i) contact mixing patterns between an infected individual and his or her contacts, (ii) age-specific susceptibility to infection, (iii) region-based behavioral susceptibility, and (iv) a potential seasonal forcing.

We incorporate age- and region-specific contact patterns between individuals, represented by contact rate between an infected individual in age-group *i*, region-group *l* and each of their contacts with susceptible in age-group *j*,region-group *k*, for different locations: at home, at work and during leisure, for each day *t*denoted by 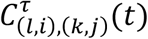, such that i*τ* ∈ {*Home, Work, Leisure*}, is the location index of the contact location index. The contact matrix 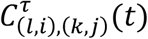 is detailed in section

### 2.1 Contact mixing patterns

We distinguish between in-home versus out-of-home transmission. Consistent with a previous study [21], we assume the in-home transmission to be fixed and independent of age, *β*_*Home*_. (See Section 2.3 Epidemiological parameters). To account for the reduced probability of infection in house following a recovery of other house members, we multiple the susceptibility inside household, *β*_*Home*_, by decay function 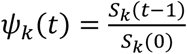. This function serve as an unbiased estimator to the proportion of susceptible individuals in the house Age-specific susceptibility rate for individuals out-of-home *β*_*j*_, was parameterized by calibrating our model with daily COVID-19 records (See Section 3. calibrated parameters).

To account for behavioral susceptibility, we explicitly considered in our model a parameter reflecting the order to maintain physical distancing, *κ*_*p*_, as vast number of countries, including Israel, adopted measures such as physical-distancing to control the susceptibility of SARS-CoV-2 [22]. This parameter was calibrated to the epidemiological data of COVID-19 in Israel. Moreover, the high regional variations in susceptibility were parameterized based on fertility rates and socioeconomic characteristics relative to the national average, using the data from Central Bureau of Statistics (CBS), *α*_*k*_. Specifically, we computed for each region the relative reduction in travels >1.5 km compared to routine *M*_*j,k,q*_(See Section 2.2 Relative reduction in travels). Our analysis indicated that for regions of low SES the change was lower, which was reflected by our model with higher susceptibility.

Seasonal patterns have been observed in common circulating HCoVs, mostly causing infections in humans between December and May in the Northern Hemisphere [23]. The two human coronaviruses 229 E and OC43 show distinct winter seasonality. In addition, many coronaviruses in animals do exhibit a distinct seasonal pattern of incidence in their natural hosts [24]. There is growing evidence that SARS-CoV-2 is also seasonal, with the optimal setting for transmission in Israel during winter [25,26]. Thus, we considered in our base-case seasonal forcing by including general seasonal variation in the susceptibility rate of the model as

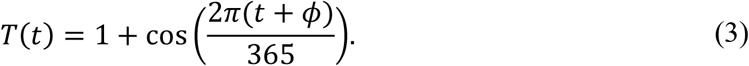

in which *φ* is seasonal offset. This formulation was previously shown to capture the seasonal variations of several respiratory infections including RSV and influenza [10,27]. We incorporated possible values of *φ* to reflect peak from December thru February (See Section 2.3 Epidemiological parameters).

Taken together, the force of infection *λ_j,k,q_*(*t*) is given by

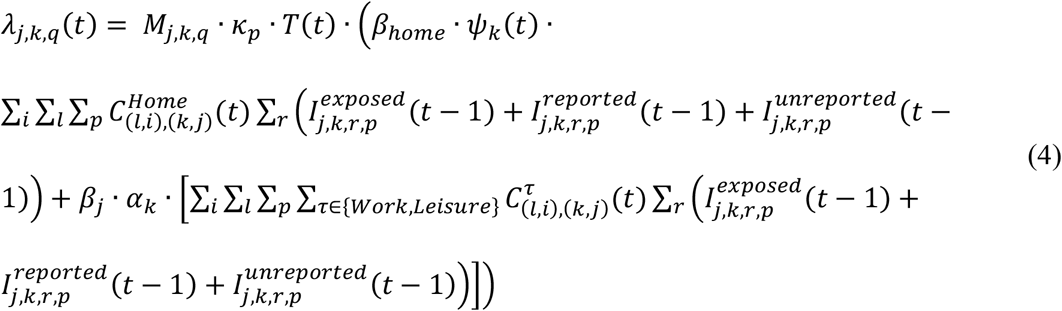

## 2. Fixed parameters

### 2.1. Contact mixing patterns

At the core of the transmission model lies the contact mixing patterns between a susceptible individual and infectious individual 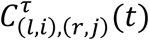. Similar to a previous study [21], the contact matrices depends on the age-group and region of residency for the susceptible individual (*l, i*), the age group and region of residency for an infectious individual (*r, j*) at location *τ* ∈ {*Home, Work, Leisure*} on day *t*. Here we detail the process of how we conducted the contact-mixing.

### Household contacts

We estimated the contact mixing at home for each region based on the average household size and its age distribution from the Israeli Central Bureau of Statistics (CBS) [28,29]. We assume all individuals in the same household will meet with each other daily regardless of the control measures applied by the country (e.g. lockdowns). The CBS data suggest that low socioeconomic status is characterized by larger and younger household size.

### Work and leisure contact patterns

#### Age-specific contacts

We parametrized the age-specific contact rates using data from a survey of daily contacts collected in eight European countries [30]. This contact data includes contact rates for different locations: works (or school for children <10), leisure. In addition, the data exhibits frequent mixing between similar age-groups, moderate mixing between children and adults in their thirties (likely their parents), and infrequent mixing between other groups. To generate the age-specific contact mixing used in our model, we used the means of each age-group over the eight countries. To ensure the matrices is symmetric and convert between age-groups used in the survey to those used in out model, we adjusted the contact matrices according to the means for reciprocal age group pairing [10].

#### Origin-destination from mobility data

Our data includes mobility records based on cellular data of >3 million users from one of the largest telecommunication companies in Israel. The data specifies movement patterns within and between 2,630 zones covering Israel, on an hourly basis, from February 1, 2020, and until May 16, 2020. To ensure privacy, if in a given hour less than 50 individuals are identified in the zone, the number of reported individuals is set to zero. We determined the location of individuals based on the triangulation of cell towers, which was found accurate to 300 meters in most cases but varied to 1 km in less populated areas. We defined users as residents of a zone based on location in which they had the highest number of signals on most nights during February 2020.

We used this data to develop aggregated origin-destination (OD) matrices between and within zones. To refrain from signal noises and identify stay points, we track only locations where users stayed for at least 15 minutes within a distance threshold of 1.7 km. The OD matrices serve as a proxy to the flow from each region to another.

Next, we integrated data from the Central Bureau of Statistics (CBS) that specifies for each zone several socioeconomic characteristics, including population size, household size, age distribution, socioeconomic score, and dominant religion. Each zone includes ∼3,500 residents. For each zone, we scaled the number of resident users of the telecommunication company to match with the actual number of residents in the zone, as recorded by the Israeli CBS. Grouping the zones by SES, and scaling for each zone the daily number of travels to one, we created an origin-destination traveling probability matrix. We found that the population is clustered, such that people of specific SES are more likely to travel to zones of the same SES during routine and even more likely during movement restrictions. These findings remain consistent when partitioning the population into resolution of 10 socioeconomic clusters, comprising the different SESs. Additionally, a similar phenomenon is observed when partitioning the population by Religious Affiliations to Arab, orthodox and non-orthodox Jewish, and also for the combination of both religious affiliation and socioeconomic clusters (Figs. S1 and S2).

**Fig. S1.**
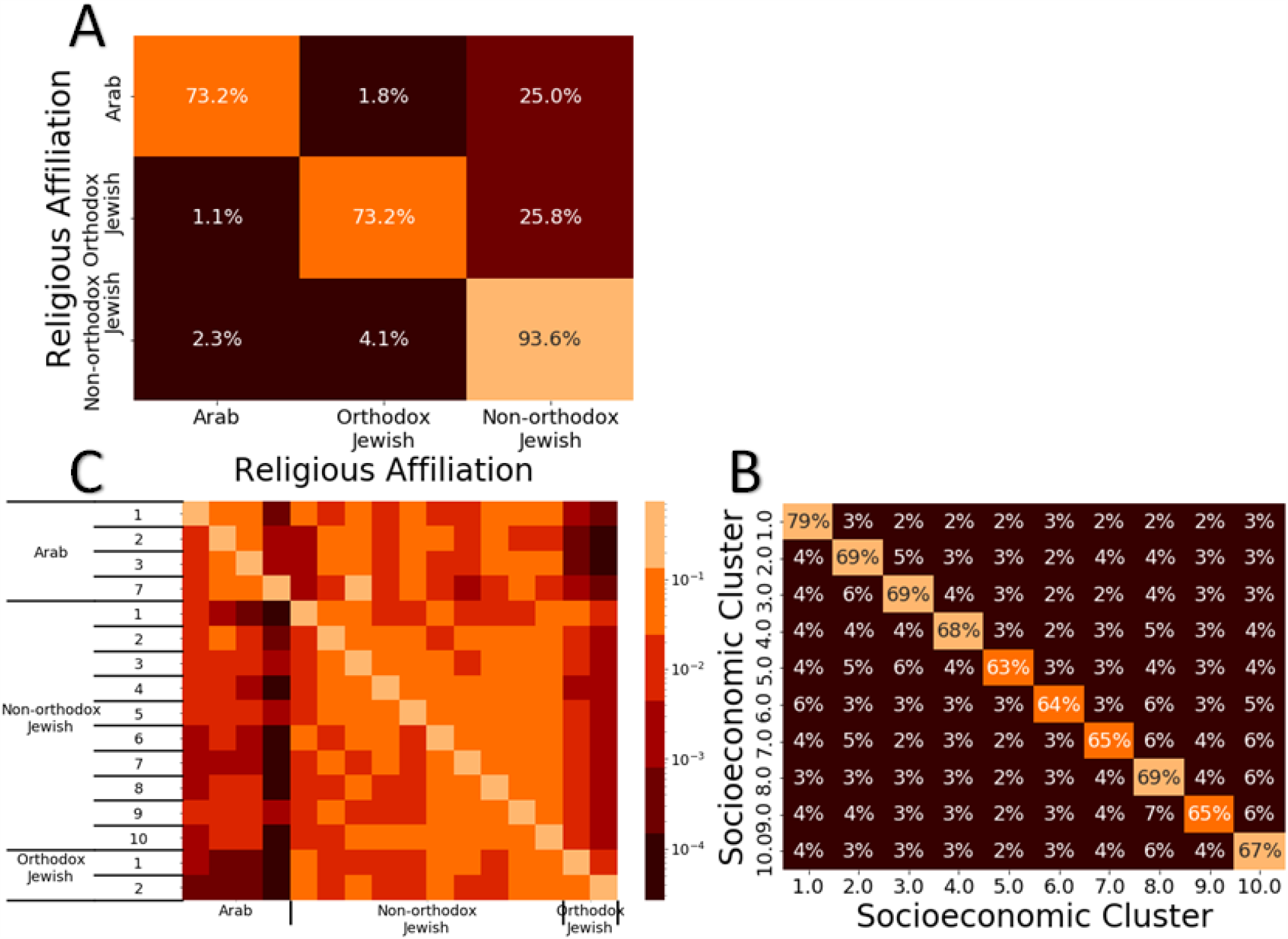
Traveling patterns during routine. Traveling patterns during February 2-29 based on (A) religious affiliation, (B) socioeconomic status, and (C) religious affiliation and socioeconomic status.

**Fig. S2.**
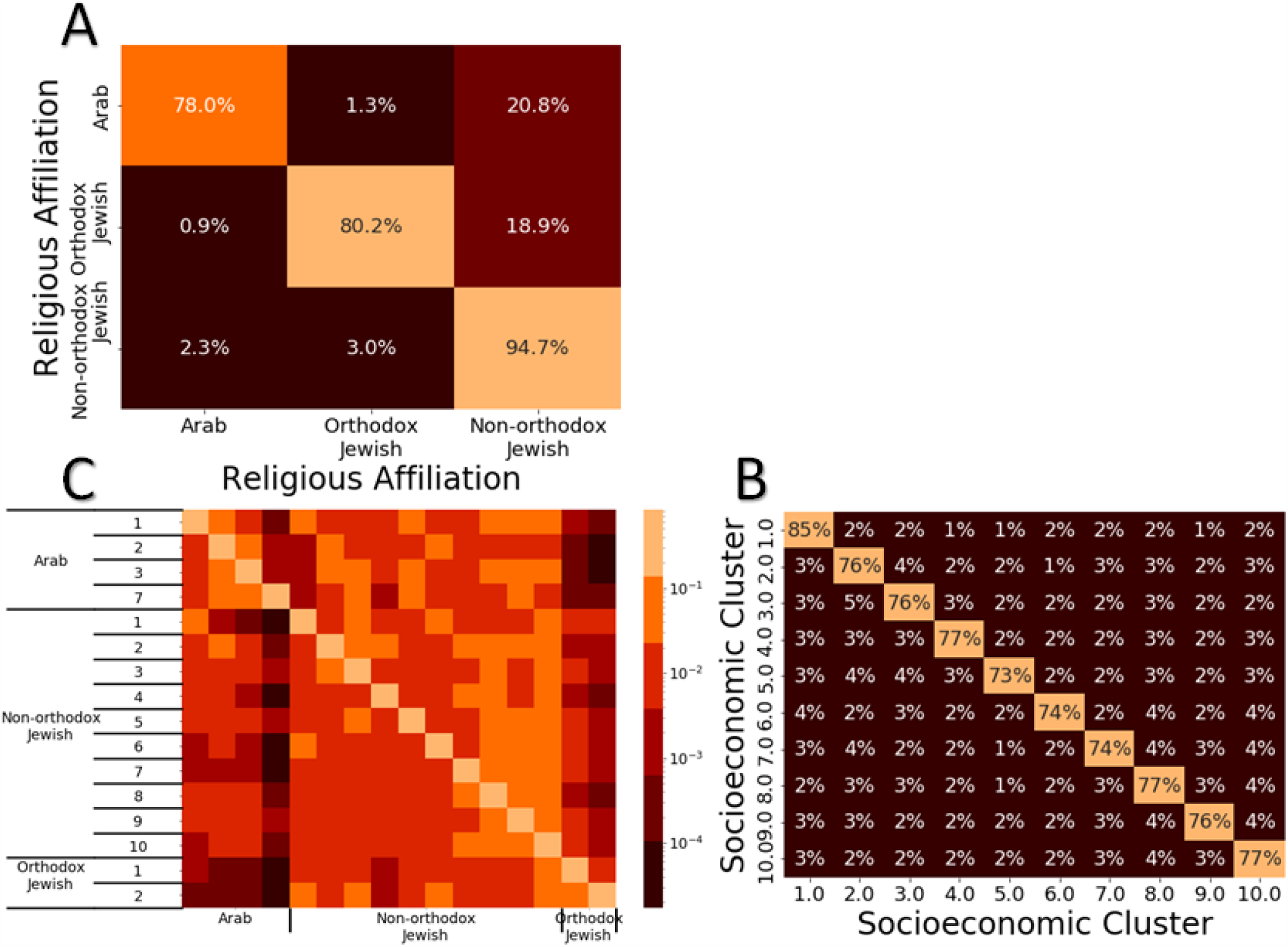
Traveling patterns during COVID-19 outbreak. Traveling patterns during March 26-April 18 based on (A) religious affiliation, (B) socioeconomic status, and (C) religious affiliation and socioeconomic status.

We used this data to develop two aggregated origin-destination (OD) matrices between and within regions from during work time 08:01-17:00 and leisure time 17:01-23:00. To incorporate the time depended travels following restrictions periods and routine we developed the two OD for the following periods: February 21 – March 13, March 14 – March 16, March 17 – March 25, March 26 – April 2, April 3 – April 6, April 7 – April 16, April 17 – May 4, May 5 – May 11.

To integrate the age-specific contact matrices and the OD matrices we multiplied the number of contacts for each age-group by the travel distribution for each region in the OD matrices. We assumed that at work, children at the age of 0-9 years old, remains at their home region. We also assumed that at leisure time children at the age of 0-9 years old movement patterns are like their parents.

### 2.2. Relative reduction in travels

For each region, we computed the relative reduction in travels >1.5 km *M*_*j,k,q*_. This measure was done scaling the daily proportion of travels more than 1.5 km out-of-home.

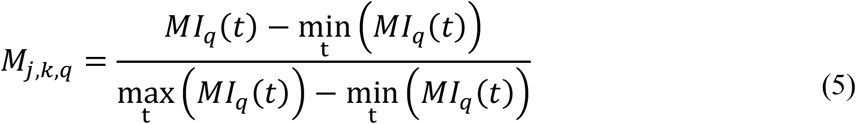

To compute this minimal and maximal values and refrain from outliers, we averaged the three minimal and three maximal values. This measure was found to be highly correlative with disease growth factor ranging between 79.2-82.8% (p value<0.001) for a shift of 12-14 days (Fig. S3). Thus, we incorporated for each region this measure in the model.

**Fig. S3.**
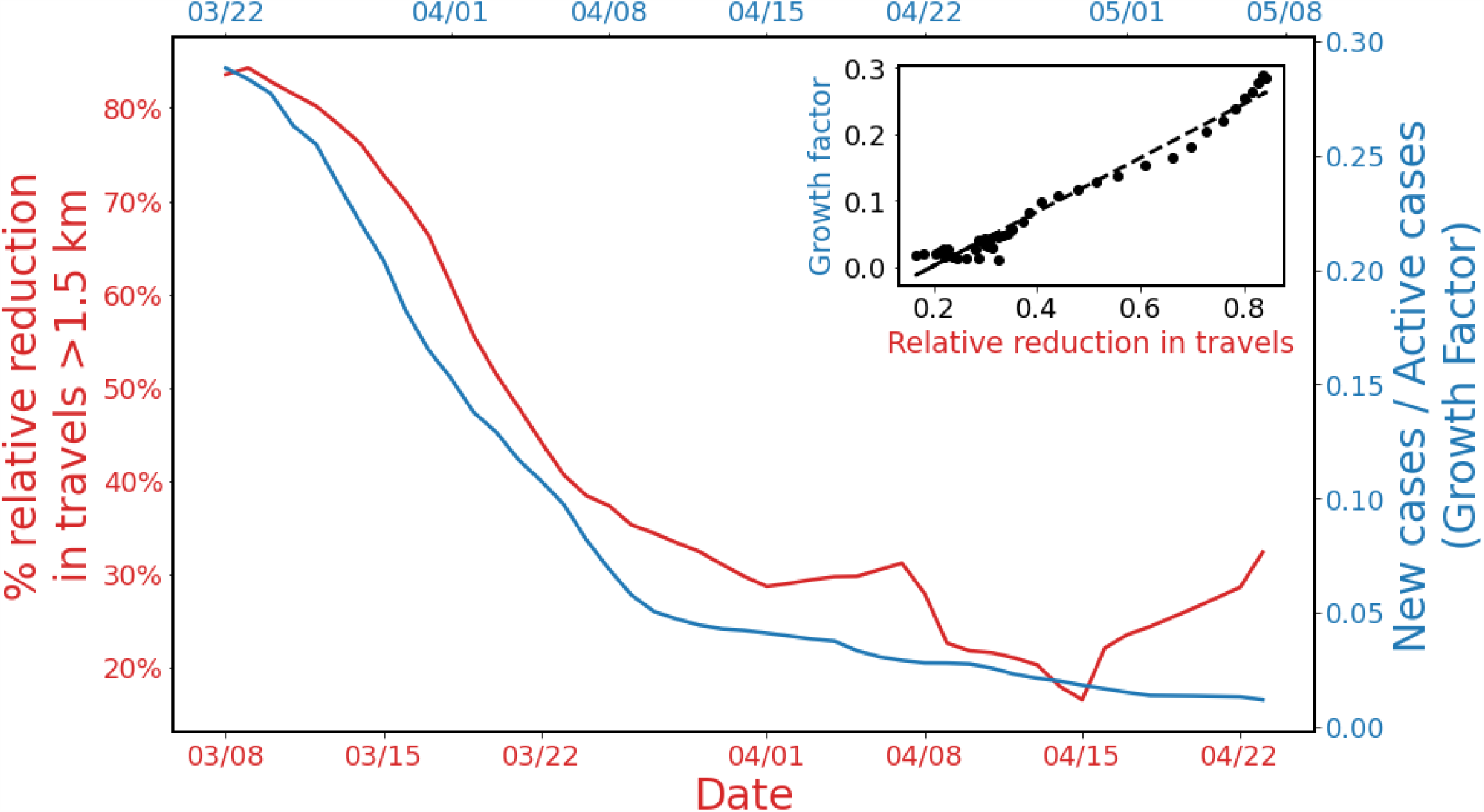
Mobility ahead of transmission. Percentage relative reduction in travels from home between March 8 and April 22 (red) and new cases per active cases between March 22 and May 8 (blue). Both plots show the weekly average. The correlation between the two is 97.0% (inserted graph).

### 2.3. Epidemiological parameters

#### Unreported cases

Under reporting arises from asymptomatic cases or mild cases of individuals that do not seek for care. The severity of SARS-Cov-2 infection is associated with age- and risk-group [31]. In addition, underreporting is affected by testing policy and testing-capabilities for each country, as well as the tendency of individuals to seek for care once clinical symptoms appear. PCR or serological screenings have yet to be conducted in Israel. Thus, we evaluated unreported cases based on PCR and serological screenings from the Czech Republic, Denmark, and Santa Clara, California, and Iceland. Similarly, to Israel, as to May 14^th^, 2020 these countries are characterized with high rates of testing and low number of severe cases. In addition, hospitals were not overwhelmed. Serological screenings from the Czech Republic suggested that each reported case corresponds to ∼5.5 unreported cases [18,20], whereas estimates from Santa Clara suggested at least 14 unreported cases for each single reported case [17]. Taken together we chose to present estimates of unreported ratios 1:5.5 (Scenario A), 1:9 (Scenario B), and 1:14 (Scenario C). It is not clear how much reutilizing antibodies are sufficient to ensure protection, and thus it is possible serological screenings serve as over estimation to determine exposure. Thus, to determine the robustness of our findings, we also considered an extreme scenario of 1:2 (Scenario D).

We estimated the proportion of under reporting for each age-group by scaling the estimates from Santa-Clara Study to the age reported cases in this region [32]. This analysis suggested that younger age-groups are more likely to be unreported. Conservatively, we assumed that all cases among individuals at high-risk are reported. Using these estimates and based on the reported cases in Israel between February 20^th^ - May 14^th^, 2020, we obtained that overall proportion of unreported cases is 85% for scenario A, 89% for scenario B, 93% for scenario C and 69% for scenario D.

**Table S1.** proportion of unreported cases. proportion of unreported cases among individuals at high risk and low risk stratified by age and overall reported cases based on the reported cases observed in Israel between February 20 and May 14, 2020.

| Scenario | Risk \ Age | 0-19 | 20-64 | ≥65 |
| --- | --- | --- | --- | --- |
| **A** | **Low** | 0.97 | 0.85 | 0.68 |
|  | **High** | 0.97 | 0.85 | 0.68 |
|  | **Total** | 0.85 |  |  |
| **B** | **Low** | 0.95 | 0.89 | 0.80 |
|  | **High** | 0.95 | 0.89 | 0.80 |
|  | **Total** | 0.89 |  |  |
| **C** | **Low** | 0.99 | 0.93 | 0.84 |
|  | **High** | 0.99 | 0.93 | 0.84 |
|  | **Total** | 0.93 |  |  |
| **D** | **Low** | 0.92 | 0.67 | 0.43 |
|  | **High** | 0.92 | 0.67 | 0.43 |
|  | **Total** | 0.69 |  |  |

#### Case fatality

The probability of death for each age-and risk-group given a reported case was evaluated based on the Israeli Ministry of Health case report data (Table S2).

**Table S2.** Probability of death for each age-and risk-group given a reported case.

| Age-group | Risk-group | Base-case value | Distribution |
| --- | --- | --- | --- |
| 0-19 | High | 0 |  |
| 20-59 | High | 0.89% | *Beta*(4,410) |
| 60-69 | High | 1.48% | *Beta*(5,312) |
| $\geq 70$ | High | 12.03% | *Beta*(52,378) |
| 0-19 | Low | 0 |  |
| 20-59 | Low | 0.06% | *Beta*(5,7759) |
| 60-69 | Low | 1.06% | *Beta*(11,995) |
| $\geq 70$ | Low | 11.33% | *Beta*(95,741) |

#### Initial morbidity(aboard)

The initial morbidity in Israel was imported by 491 citizens who returned from overseas. The first infected traveler identified on February 20, and by March 9^th^, 2020 a self-quarantine was mandatory for all returning. Most of the flights to Israel arrive from the developed countries. Thus, we distributed the these cases in each day of the 18 days proportionally to the daily new cases in Italy, which had the hardest hit among developed countries [33]. To account for under reporting, we multiplied the number of cases in each day according to the unreported scenarios we considered (Table S1). We entered these initial spreaders, *ε*_*j,k,r,i*_(*t*), to the exposed compartment.

#### Susceptibility at-home

We distinguish between in-home versus out-of-home transmission. Consistent with a previous study [21]. We specifically distinguish between the susceptibility of those settings. We estimated the in-home susceptibility rate, *β*_*home*_, based on a previous study that showed a secondary attack rate of 16.3% throughout the entire infectious period [34].

**Table S3.**
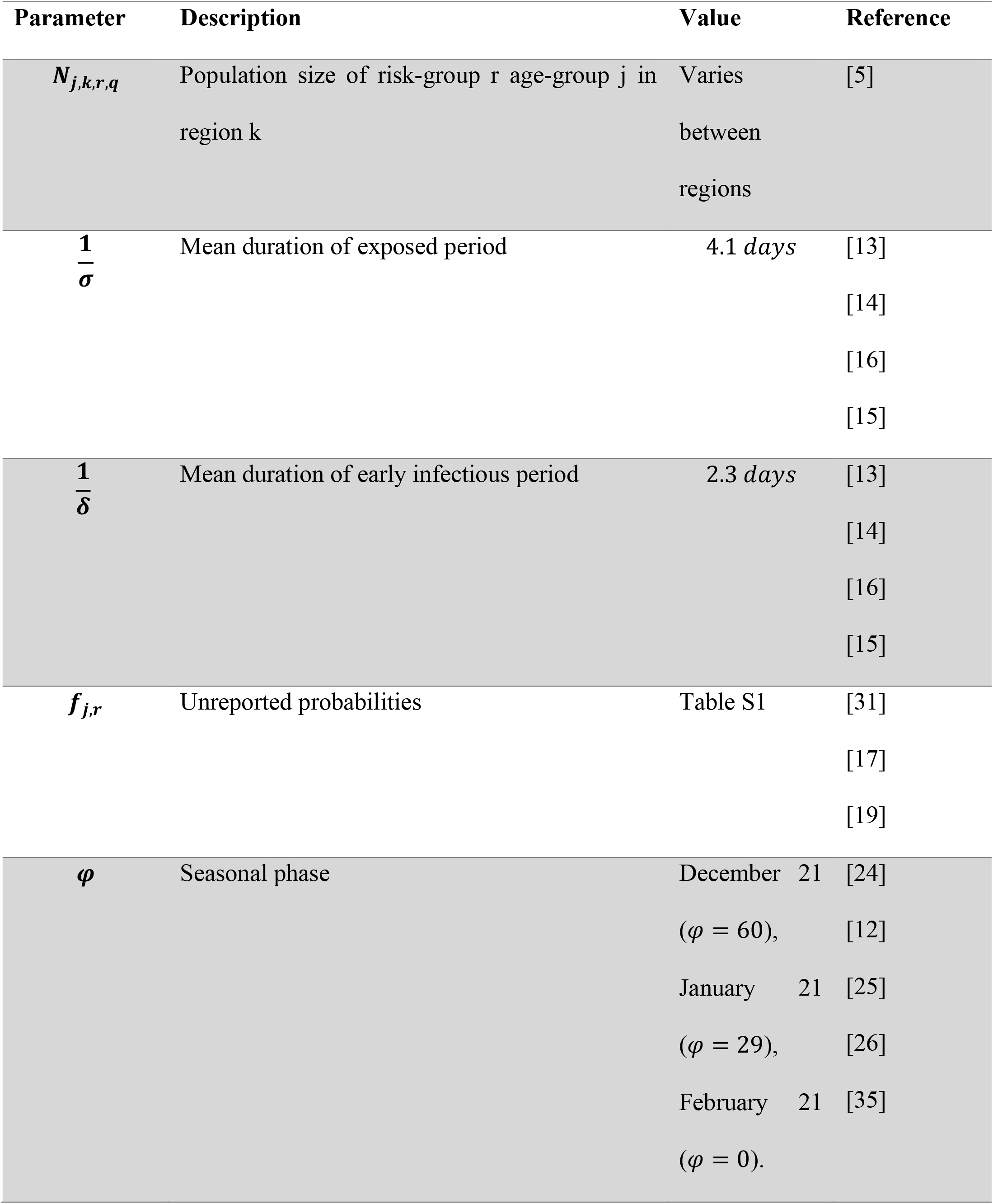
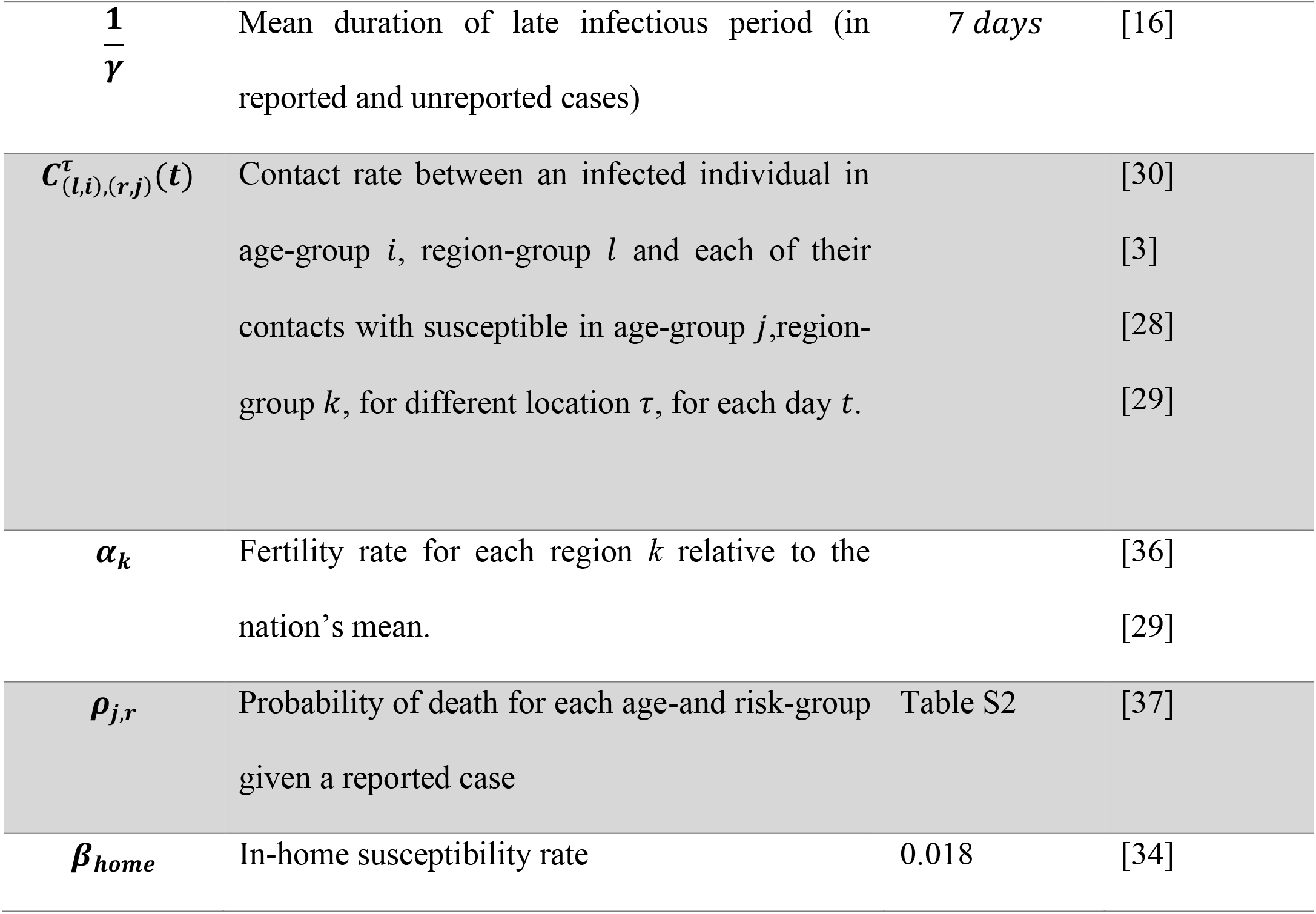
Fixed parameters used in the transmission model.

| Parameter | Description | Value | Reference |
| --- | --- | --- | --- |
| $N_{j,k,r,q}$ | Population size of risk-group r age-group j in region k | Varies between regions | [5] |
| $\frac{1}{\sigma}$ | Mean duration of exposed period | 4.1 days | [13] |
|  |  |  | [14] |
|  |  |  | [16] |
|  |  |  | [15] |
| $\frac{1}{\delta}$ | Mean duration of early infectious period | 2.3 days | [13] |
|  |  |  | [14] |
|  |  |  | [16] |
|  |  |  | [15] |
| $f_{j,r}$ | Unreported probabilities | Table S1 | [31] |
|  |  |  | [17] |
|  |  |  | [19] |
| $\varphi$ | Seasonal phase | December 21 | [24] |
| | | ($\varphi = 60$), | [12] |
|  |  | January 21 | [25] |
| | | ($\varphi = 29$), | [26] |
|  |  | February 21 | [35] |
| | | ($\varphi = 0$). | |
| $\frac{1}{\gamma}$ | Mean duration of late infectious period (in reported and unreported cases) | 7 days | [16] |
| $C_{(l,i),(r,j)}^{\tau}(t)$ | Contact rate between an infected individual in age-group $i$, region-group $l$ and each of their contacts with susceptible in age-group $j$, region-group $k$, for different location $\tau$, for each day $t$. | | [30]<br>[3]<br>[28]<br>[29] |
| $\alpha_k$ | Fertility rate for each region $k$ relative to the nation's mean. | | [36]<br>[29] |
| $\rho_{j,r}$ | Probability of death for each age-and risk-group given a reported case | Table S2 | [37] |
| $\beta_{home}$ | In-home susceptibility rate | 0.018 | [34] |

## 3. Calibrated parameters

To estimate empirically unknown epidemiological parameters, we calibrated our model to daily age-stratified cases of COVID-19 confirmed by PCR tests in 30 subdistricts covering Israel between March 1 until May 10. We shifted the data 11 days backward, to compensate for the lag between the date of infection and the date of first positive SARS-CoV2 test result, which was found to be 10.5 days on average according to MOH’s epidemiological investigations. We applied a central moving average with window of three days before and after the data point, on the data to reduce noise caused by weekly patterns.

The calibration was conducted on a 30-subdistrict level rather than 250 regions to ensure there are sufficient time-series data points in each location for each age group. The stratification is based on the 16 formal districts, which we further stratified such that the sub districts will be homogenous in terms of their SES and religious affiliation (Table S4). To calibrate the model to the incidence data, we maximized the likelihood assuming a normal distribution of the error between model predictions and incidence data. This was achieved by using the truncated Newton (TNC) algorithm. We calibrated the model for 16 different scenarios of unreported cases and seasonal forcing. The final transmission model included five parameters without constraints imposed from previous data: reduced susceptibility due to physical distancing *κ*_*p*_, and susceptibility rate based on age-groups *j*: 0-19, 20-39, 40-59, and >60 (Table S5).

We used an F-test of equality of variances to compare between models 1) with vs. without consideration of seasonal forcing, 2) with and without consideration of human mobility, 3) with and without consideration of regional fertility. We denote that in all three comparisons, the number of calibrated parameters is constant and equal to five. Our tests suggested that models that do not include the mobility data (p.value<0.01), and the regional fertilities (p.value<0.01) were significantly worse. We also found that models that accounted for seasonal forcing yielded higher, but not significant (p value<0.35), likelihood than models that did not account for the seasonal forcing.

**Table S4.** 30 subdistricts calibrated.

| **Sub-district number** | **Name** | **Population Size** |
| --- | --- | --- |
| 1 | Jerusalem and sub. | 778,503 |
| 2 | Bet Shemesh | 120,164 |
| 3 | Jerusalem and sub. (Orthodox Jewish) | 265,313 |
| 4 | Zefat | 138,618 |
| 5 | Zefat (Israeli Arabs) | 23,772 |
| 6 | Kinneret (Jewish) | 98,178 |
| 7 | Jezreel Valley (Israeli Arabs) | 159,112 |
| 8 | Jezreel Valley (Jewish) | 351,446 |
| 9 | Akko (Israeli Arabs) | 357,341 |
| 10 | Akko (Jewish) | 314,607 |
| 11 | Ramat Hagolan | 51,980 |
| 12 | Haifa (Israeli Arabs) | 35,637 |
| 13 | Haifa (Jewish) | 589,951 |
| 14 | Hadera (Israeli Arabs) | 115,000 |
| 15 | Hadera (Jewish) | 315,593 |
| 16 | Sharon (Israeli Arabs) | 85,729 |
| 17 | Sharon (Jewish) | 412,638 |
| 18 | Petah Tiqwa (Israeli Arabs) | 27,455 |
| 19 | Petah Tiqwa (Orthodox Jewish) | 49,549 |
| 20 | Petah Tiqwa (Secular Jewish) | 680,836 |
| 21 | Ramla | 323,352 |
| 22 | Rehovot | 661,079 |
| 23 | Tel Aviv – Yafo | 820,271 |
| 24 | Bnei Brak | 211,259 |
| 25 | Tel Aviv suburbs | 464,974 |
| 26 | Ashqelon | 559,556 |
| 27 | Beer Sheva (Israeli Arabs) | 196,311 |
| 28 | Beer Sheva (Jewish) | 504,831 |
| 29 | Judea and Samaria | 267,832 |
| 30 | Judea and Samaria (Orthodox Jewish) | 155,095 |

**Table S5.** Calibrated parameters.

| Model configuration | Seasonal forcing peak | Unreported [%] | Physical distancing Coefficient $\kappa_{physical}$ | Susceptibility among age-group 0-19[y] | Susceptibility among age-group 20-39[y] | Susceptibility among age-group 40-59[y] | Susceptibility among age-group 60+[y] | Likelihood of calibration to data $-\log(l)$ |
| --- | --- | --- | --- | --- | --- | --- | --- | --- |
| Full model | No-seasonality | 69 | 0.248 | 0.094 | 0.054 | 0.042 | 0.311 | -25.766 |
| Full model | No-seasonality | 85 | 0.232 | 0.119 | 0.053 | 0.052 | 0.166 | -25.743 |
| Full model | No-seasonality | 89 | 0.234 | 0.057 | 0.076 | 0.047 | 0.116 | -25.494 |
| Full model | No-seasonality | 93 | 0.246 | 0.119 | 0.036 | 0.054 | 0.184 | -25.876 |
| Full model | December 21 | 69 | 0.272 | 0.038 | 0.023 | 0.020 | 0.128 | -25.856 |
| Full model | December 21 | 85 | 0.306 | 0.044 | 0.021 | 0.024 | 0.109 | -25.862 |
| Full model | December 21 | 89 | 0.355 | 0.025 | 0.021 | 0.025 | 0.144 | -25.998 |
| Full model | December 21 | 93 | 0.274 | 0.058 | 0.015 | 0.023 | 0.083 | -25.917 |
| Full model | January 21 | 69 | 0.364 | 0.043 | 0.025 | 0.024 | 0.151 | -25.835 |
| Full model | January 21 | 85 | 0.420 | 0.049 | 0.027 | 0.024 | 0.148 | -25.787 |
| Full model | January 21 | 89 | 0.349 | 0.035 | 0.031 | 0.029 | 0.167 | -25.957 |
| Full model | January 21 | 93 | 0.295 | 0.059 | 0.020 | 0.030 | 0.112 | -25.898 |
| Full model | February 21 | 69 | 0.347 | 0.063 | 0.039 | 0.033 | 0.248 | -25.822 |
| Full model | February 21 | 85 | 0.464 | 0.051 | 0.036 | 0.034 | 0.199 | -25.813 |
| Full model | February 21 | 89 | 0.417 | 0.052 | 0.045 | 0.041 | 0.229 | -25.916 |
| Full model | February 21 | 93 | 0.411 | 0.100 | 0.030 | 0.034 | 0.157 | -25.827 |
| Without mobility | January 21 | 85 | 0.127 | 0.022 | 0.035 | 0.022 | 0.162 | -25.129 |
| Without mobility | January 21 | 89 | 0.133 | 0.031 | 0.029 | 0.022 | 0.133 | -25.206 |
| Without mobility | January 21 | 93 | 0.098 | 0.049 | 0.030 | 0.023 | 0.121 | -25.139 |
| Without fertility | January 21 | 85 | 0.633 | 0.056 | 0.027 | 0.018 | 0.013 | -25.311 |

## 4. Further simulation results

We found that a global lockdown strategy had a larger temporal effect than local lockdowns and had by greater oscillations (Fig. S4). We present here a model with seasonal forcing. Our model projections suggested that global lockdowns were less efficient and effective compared to a strategy that targets locally the elderly. However, due to high variability between the 250 regions considered, some regions undergo multiple lockdowns, while others will not undergo lockdowns. Local lockdowns that specifically target children decreases the local morbidity, but in the long run increases mortality, while lockdowns of individuals at high-risk has a moderate impact on transmission but decreases mortality.

These findings where robust across all settings considered (Tables S3 and S5), when we accounted for seasonal forcing (Main text, Figs. 4 and 5), and without seasonal forcing (Fig. S5).

**Fig. S4.**
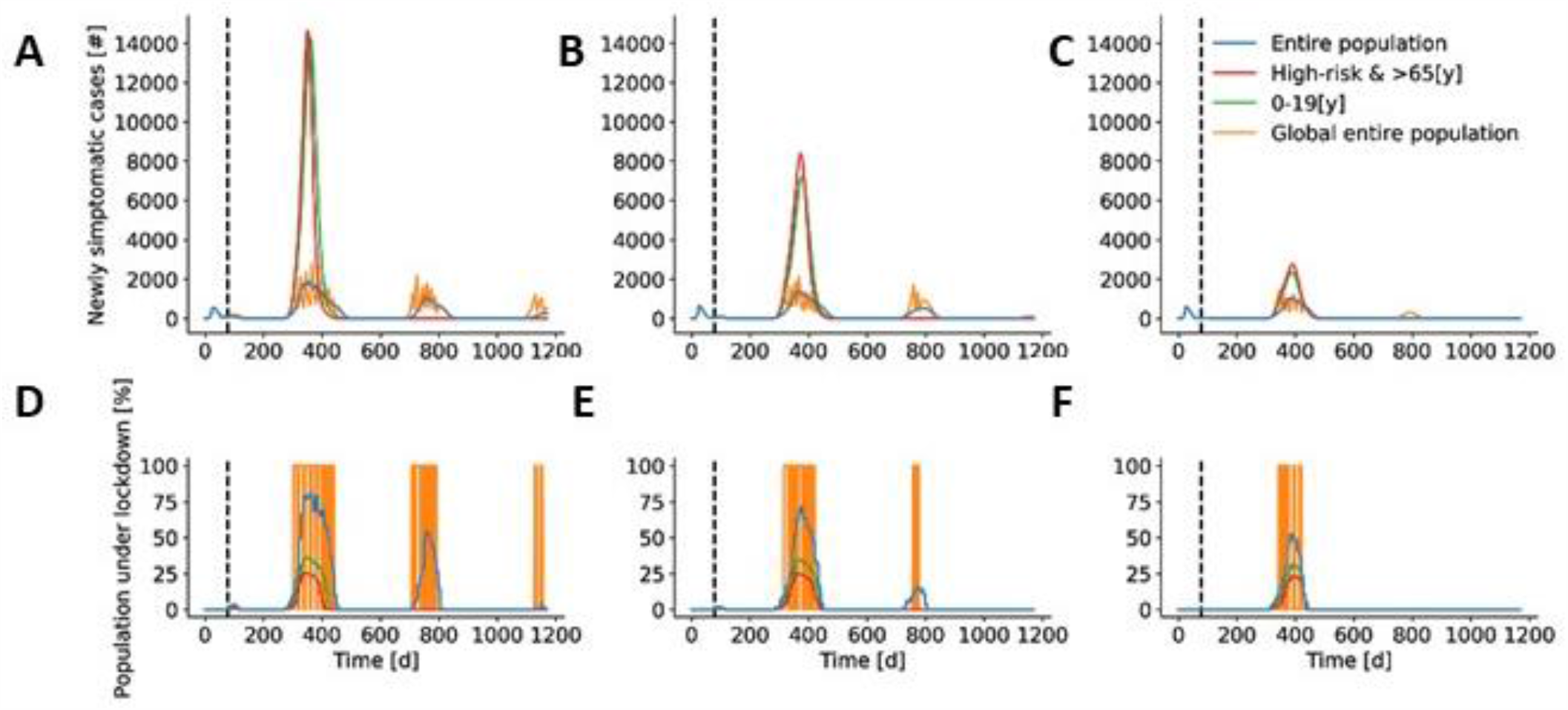
Model demonstration for a threshold of 1 per 10000 for the lockdown strategies with seasonal forcing peaking on January 21. (A – C) projected daily new reported cases for different lockdown strategies. (D – F) Projected daily percentage of population under lockdown. (A, D) for a unreported cases of 85%. (B, E) for 89%, and (C, F) for 93%.

**Fig. S5.**
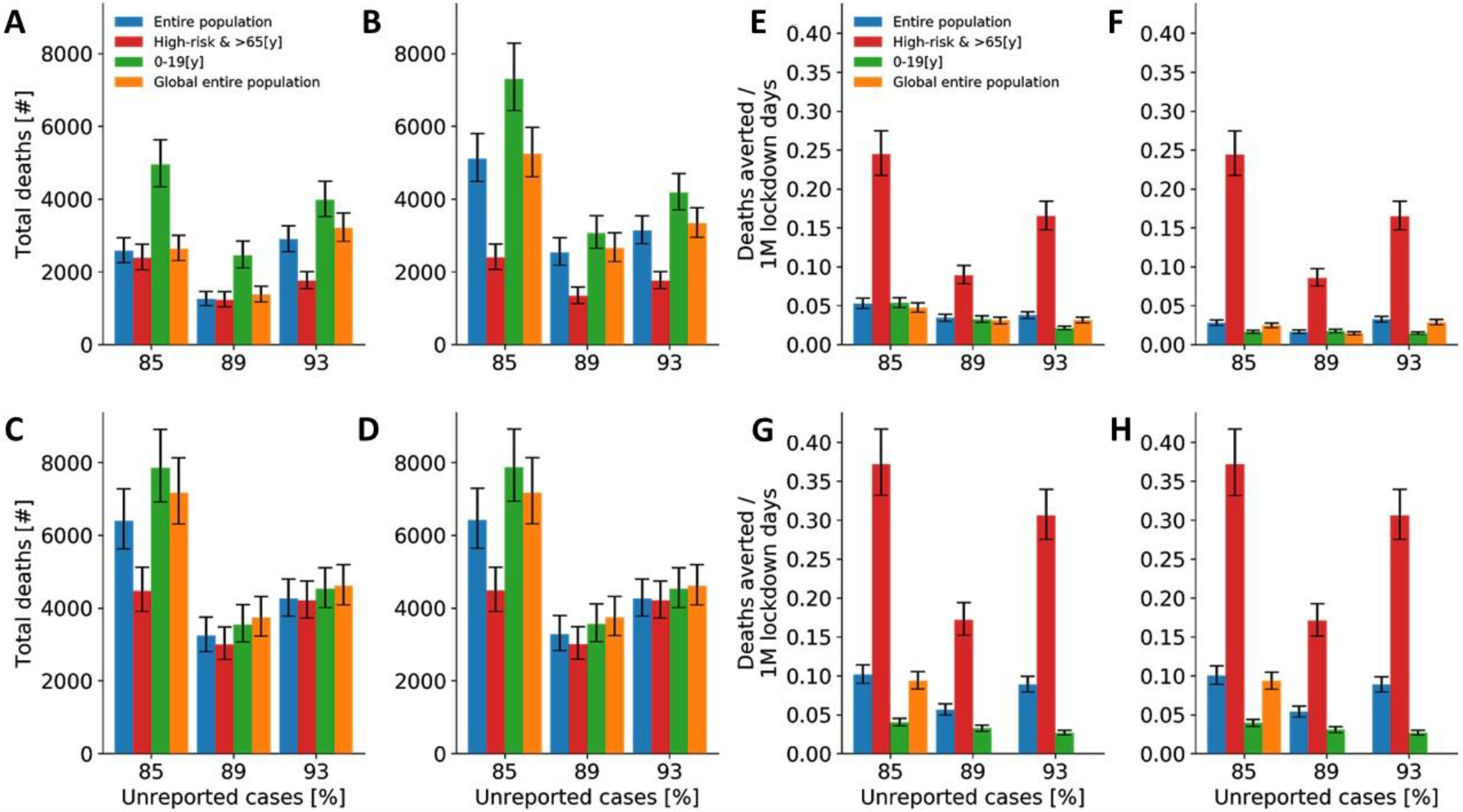
Effectiveness and efficiency of temporal-local lockdowns without seasonal forcing. Median and interquartile values of model projections after implementation of strategies (A, C, E, G) after one year and (B, D, F, H) after three years. (A, B, E, F) The thresholds for lockdowns in a local region are 1/10,000 [cases/individuals] and (C, D, G, H) 5/10000 [cases/individuals]. Effectiveness (A – D), efficiency (E – G).

## References

1. Coronavirus disease 2019. https://www.who.int/emergencies/diseases/novel-coronavirus-2019?gclid=EAIaIQobChMIm_vn_6XN6QIVhPdRCh0CbQI_EAAYASAAEgIa7PD_BwE. Accessed 24 May 2020.

2. Trip.com COVID-19 Country/Region Entry Restrictions. https://www.trip.com/travel-restrictions-covid-19/. Accessed 24 May 2020.

3. The Novel Coronavirus - Israel Ministry of Health. https://govextra.gov.il/ministry-of-health/corona/corona-virus-en/. Accessed 24 May 2020.

4. Fernandes N. Economic Effects of Coronavirus Outbreak (COVID-19) on the World Economy. SSRN Electron J. 2020.

5. PsyArXiv Preprints | Students under lockdown: Assessing change in students’ social networks and mental health during the COVID-19 crisis. https://psyarxiv.com/ua6tq. Accessed 8 Jun 2020.

6. Kazmi SSH, Hasan K, Talib S, Saxena S. COVID-19 and Lockdwon: A Study on the Impact on Mental Health. SSRN Electron J. 2020.

7. Armitage R, Nellums LB. COVID-19 and the consequences of isolating the elderly. 2020. doi:10.1016/S2468-2667(20)30061-X.

8. Ahmed F, Ahmed N, Pissarides C, Stiglitz J. Why inequality could spread COVID-19. The Lancet Public Health. 2020;5:e240.

9. Charu V, Zeger S, Gog J, Bjørnstad ON, Kissler S, Simonsen L, et al. Human mobility and the spatial transmission of influenza in the United States. PLOS Comput Biol. 2017;13:e1005382. doi:10.1371/journal.pcbi.1005382.

10. Yamin D, Gavious A, Solnik E, Davidovitch N, Balicer RD, Galvani AP, et al. An Innovative Influenza Vaccination Policy: Targeting Last Season’s Patients. PLoS Comput Biol. 2014.

11. Prem K, Liu Y, Russell TW, Kucharski AJ, Eggo RM, Davies N, et al. The effect of control strategies to reduce social mixing on outcomes of the COVID-19 epidemic in Wuhan, China: a modelling study. Lancet Public Heal. 2020;5:e261–70.

12. Finger F, Genolet T, Mari L, Constantin De Magny G, Magloire Manga N, Rinaldo A, et al. Mobile phone data highlights the role of mass gatherings in the spreading of cholera outbreaks. PNAS. 2016;113:6421–6.

13. Kraemer MUG, Yang CH, Gutierrez B, Wu CH, Klein B, Pigott DM, et al. The effect of human mobility and control measures on the COVID-19 epidemic in China. Science. 2020;368:493–7.

14. Bendavid E, Mulaney B, Sood N, Shah S, Ling E, Bromley-Dulfano R, et al. COVID-19 Antibody Seroprevalence in Santa Clara County, California. medRxiv. 2020;:2020.04.14.20062463. doi:10.1101/2020.04.14.20062463.

15. Chow N, Fleming-Dutra K, Gierke R, Hall A, Hughes M, Pilishvili T, et al. Preliminary estimates of the prevalence of selected underlying health conditions among patients with coronavirus disease 2019 - United States, February 12-March 28, 2020. Morbidity and Mortality Weekly Report. 2020;69:382–6.

16. Brodin P. Why is COVID-19 so mild in children? Acta Paediatr. 2020;109:1082–3. doi:10.1111/apa.15271.

17. Dr. Anthony Fauci On U.S Efforts To Develop A Coronavirus Vaccine?: NPR. https://www.npr.org/2020/05/22/860682211/dr-anthony-fauci-on-u-s-efforts-to-develop-a-coronavirus-vaccine. Accessed 25 May 2020.

18. Zhou F, Yu T, Du R, Fan G, Liu Y, Liu Z, et al. Clinical course and risk factors for mortality of adult inpatients with COVID-19 in Wuhan, China: a retrospective cohort study. Lancet. 2020;395:1054–62.

19. Li W, Zhang B, Lu J, Liu S, Chang Z, Cao P, et al. The characteristics of household transmission of COVID-19. doi:10.1093/cid/ciaa450/5821281.

20. Vynnycky E, White R. Introduction. The basics: infections, transmission and models. In: An Introduction to Infectious Disease Modelling. 2010.

21. Molinari NAM, Ortega-Sanchez IR, Messonnier ML, Thompson WW, Wortley PM, Weintraub E, et al. The annual impact of seasonal influenza in the US: Measuring disease burden and costs. Vaccine. 2007.

22. Fiore AE, Fry A, Shay D, Gubareva L, Bresee JS, Uyeki TM, et al. Antiviral agents for the treatment and chemoprophylaxis of influenza --- recommendations of the Advisory Committee on Immunization Practices (ACIP). MMWR Surveill Summ Morb Mortal Wkly report Surveill Summ / CDC. 2011.

23. Lauer SA, Grantz KH, Bi Q, Jones FK, Zheng Q, Meredith HR, et al. The Incubation Period of Coronavirus Disease 2019 (COVID-19) From Publicly Reported Confirmed Cases: Estimation and Application. Ann Intern Med. 2020.

24. Linton NM, Kobayashi T, Yang Y, Hayashi K, Akhmetzhanov AR, Jung S, et al. Incubation Period and Other Epidemiological Characteristics of 2019 Novel Coronavirus Infections with Right Truncation: A Statistical Analysis of Publicly Available Case Data. J Clin Med. 2020;9:538. doi:10.3390/jcm9020538.

25. Gandhi M, Yokoe DS, Havlir D V. Asymptomatic Transmission, the Achilles’ Heel of Current Strategies to Control Covid-19. N Engl J Med. 2020.

26. He X, Lau EHY, Wu P, Deng X, Wang J, Hao X, et al. Temporal dynamics in viral shedding and transmissibility of COVID-19. Nat Med. 2020;26:672–5.

27. Zou L, Ruan F, Huang M, Liang L, Huang H, Hong Z, et al. SARS-CoV-2 viral load in upper respiratory specimens of infected patients. New England Journal of Medicine. 2020;382:1177–9. doi:10.1056/NEJMc2001737.

28. Zheng S, Fan J, Yu F, Feng B, Lou B, Zou Q, et al. Viral load dynamics and disease severity in patients infected with SARS-CoV-2 in Zhejiang province, China, January-March 2020: retrospective cohort study. BMJ. 2020;369:m1443.

29. Ni L, Ye F, Cheng M-L, Feng Y, Deng Y-Q, Zhao H, et al. Detection of SARS-CoV-2-specific humoral and cellular immunity in COVID-19 convalescent individuals. Immunity. 2020. doi:10.1016/j.immuni.2020.04.023.

30. Ng OW, Chia A, Tan AT, Jadi RS, Leong HN, Bertoletti A, et al. Memory T cell responses targeting the SARS coronavirus persist up to 11 years post-infection. Vaccine. 2016;34:2008–14.

31. Yamin D, Jones FK, DeVincenzo JP, Gertler S, Kobiler O, Townsend JP, et al. Vaccination strategies against respiratory syncytial virus. Proc Natl Acad Sci U S A. 2016.

32. Kissler SM, Tedijanto C, Goldstein E, Grad YH, Lipsitch M. Projecting the transmission dynamics of SARS-CoV-2 through the postpandemic period. Science (80-). 2020;368:eabb5793.

33. Mossong J, Hens N, Jit M, Beutels P, Auranen K, Mikolajczyk R, et al. Social contacts and mixing patterns relevant to the spread of infectious diseases. PLoS Med. 2008.

34. Gaunt ER, Hardie A, Claas ECJ, Simmonds P, Templeton KE. Epidemiology and Clinical Presentations of the Four Human Coronaviruses 229E, HKU1, NL63, and OC43 Detected over 3 Years Using a Novel Multiplex Real-Time PCR Method. J Clin Microbiol. 2010;48:2940–7. doi:10.1128/JCM.00636-10.

35. Zimmermann P, Curtis N. Coronavirus infections in children including COVID-19: An overview of the epidemiology, clinical features, diagnosis, treatment and prevention options in children. Pediatric Infectious Disease Journal. 2020;39:355–68.

36. Wang J, Tang K, Feng K, Lv W. High Temperature and High Humidity Reduce the Transmission of COVID-19. SSRN Electron J. 2020.

37. Bock Axelsen J, Yaari R, Grenfell BT, Stone L. Multiannual forecasting of seasonal influenza dynamics reveals climatic and evolutionary drivers. doi:10.1073/pnas.1321656111.

38. (No Title). https://www.zva.gov.lv/sites/default/files/inline-files/05_07_covid-19-rapid-risk-assessment-coronavirus-disease-2019-ninth-update-23-april-2020-1.pdf. Accessed 30 May 2020.

39. Marcel S, Christian A, Richard N, Silvia S, Emma H, Jacques F, et al. COVID-19 epidemic in Switzerland: on the importance of testing, contact tracing and isolation.

40. Hope C. LOCKDOWN: A FIRST UTILITARIAN ANALYSIS.

41. Liu T, Wu S, Tao H, Zeng G, Zhou F, Guo F, et al. Prevalence of IgG antibodies to SARS-CoV-2 in Wuhan - implications for the ability to produce long-lasting protective antibodies against SARS-CoV-2. medRxiv. 2020;:2020.06.13.20130252. doi:10.1101/2020.06.13.20130252.

42. Wu JT, Leung K✉, Leung K, Bushman M, Kishore N, Niehus R, et al. Estimating clinical severity of COVID-19 from the transmission dynamics in Wuhan, China. doi:10.1038/s41591-020-0822-7.

43. McMichael TM, Currie DW, Clark S, Pogosjans S, Kay M, Schwartz NG, et al. Epidemiology of Covid-19 in a Long-Term Care Facility in King County, Washington. N Engl J Med. 2020;382:2005–11. doi:10.1056/NEJMoa2005412.

44. Bialek S, Gierke R, Hughes M, McNamara LA, Pilishvili T, Skoff T. Coronavirus Disease 2019 in Children — United States, February 12–April 2, 2020. MMWR Morb Mortal Wkly Rep. 2020;69:422–6. doi:10.15585/mmwr.mm6914e4.

## References

1. Vynnycky E, White R. Introduction. The basics: infections, transmission and models. An Introduction to Infectious Disease Modelling. 2010.

2. Yamin D, Gavious A, Solnik E, Davidovitch N, Balicer RD, Galvani AP, et al. An Innovative Influenza Vaccination Policy: Targeting Last Season’s Patients. PLoS Comput Biol. 2014. doi:10.1371/journal.pcbi.1003643

3. Medlock J, Galvani AP. Optimizing influenza vaccine distribution. Science (80-). 2009. doi:10.1126/science.1175570

4. Ndeffo Mbah ML, Medlock J, Meyers LA, Galvani AP, Townsend JP. Optimal targeting of seasonal influenza vaccination toward younger ages is robust to parameter uncertainty. Vaccine. 2013. doi:10.1016/j.vaccine.2013.04.052

5. Molinari NAM, Ortega-Sanchez IR, Messonnier ML, Thompson WW, Wortley PM, Weintraub E, et al. The annual impact of seasonal influenza in the US: Measuring disease burden and costs. Vaccine. 2007. doi:10.1016/j.vaccine.2007.03.046

6. Fiore AE, Fry A, Shay D, Gubareva L, Bresee JS, Uyeki TM, et al. Antiviral agents for the treatment and chemoprophylaxis of influenza --- recommendations of the Advisory Committee on Immunization Practices (ACIP). MMWR Surveill Summ Morb Mortal Wkly report Surveill Summ / CDC. 2011.

7. Ni L, Ye F, Cheng M-L, Feng Y, Deng Y-Q, Zhao H, et al. Detection of SARS-CoV-2-specific humoral and cellular immunity in COVID-19 convalescent individuals. Immunity. 2020 [cited 13 May 2020]. doi:10.1016/j.immuni.2020.04.023

8. Bao L, Deng W, Gao H, Xiao C, Liu J, Xue J, et al. Reinfection could not occur in SARS-CoV-2 infected rhesus macaques. bioRxiv. 2020; 2020.03.13.990226. doi:10.1101/2020.03.13.990226

9. Ng OW, Chia A, Tan AT, Jadi RS, Leong HN, Bertoletti A, et al. Memory T cell responses targeting the SARS coronavirus persist up to 11 years post-infection. Vaccine. 2016;34: 2008–2014. doi:10.1016/j.vaccine.2016.02.063

10. Yamin D, Jones FK, DeVincenzo JP, Gertler S, Kobiler O, Townsend JP, et al. Vaccination strategies against respiratory syncytial virus. Proc Natl Acad Sci U S A. 2016. doi:10.1073/pnas.1522597113

11. Designed Research; S APGMM. Projecting hospital utilization during the COVID-19 outbreaks in the United States. 2020;117: 9122–9126. doi:10.1073/pnas.2004064117/-/DCSupplemental

12. Kissler SM, Tedijanto C, Goldstein E, Grad YH, Lipsitch M. Supplementary Materials for Projecting the transmission dynamics of SARS-CoV-2 through the postpandemic period. doi:10.1126/science.abb5793

13. Lauer SA, Grantz KH, Bi Q, Jones FK, Zheng Q, Meredith HR, et al. The Incubation Period of Coronavirus Disease 2019 (COVID-19) From Publicly Reported Confirmed Cases: Estimation and Application. Ann Intern Med. 2020. doi:10.7326/M20-0504

14. Linton NM, Kobayashi T, Yang Y, Hayashi K, Akhmetzhanov AR, Jung S, et al. Incubation Period and Other Epidemiological Characteristics of 2019 Novel Coronavirus Infections with Right Truncation: A Statistical Analysis of Publicly Available Case Data. J Clin Med. 2020;9: 538. doi:10.3390/jcm9020538

15. Gandhi M, Yokoe DS, Havlir D V. Asymptomatic Transmission, the Achilles’ Heel of Current Strategies to Control Covid-19. N Engl J Med. 2020. doi:10.1056/nejme2009758

16. He X, Lau EHY, Wu P, Deng X, Wang J, Hao X, et al. Temporal dynamics in viral shedding and transmissibility of COVID-19. Nat Med. 2020;26: 672–675. doi:10.1038/s41591-020-0869-5

17. Bendavid E, Mulaney B, Sood N, Shah S, Ling E, Bromley-Dulfano R, et al. COVID-19 Antibody Seroprevalence in Santa Clara County, California. medRxiv. 2020; 2020.04.14.20062463. doi:10.1101/2020.04.14.20062463

18. (No Title). [cited 30 May 2020]. Available: https://www.zva.gov.lv/sites/default/files/inline-files/05_07_covid-19-rapid-risk-assessment-coronavirus-disease-2019-ninth-update-23-april-2020-1.pdf

19. Gudbjartsson DF, Helgason A, Jonsson H, Magnusson OT, Melsted P, Norddahl GL, et al. Spread of SARS-CoV-2 in the Icelandic Population. N Engl J Med. 2020. doi:10.1056/nejmoa2006100

20. Czech study shows very low COVID-19 incidence in population. [cited 30 May 2020]. Available: https://medicalxpress.com/news/2020-05-czech-covid-incidence-population.html

21. Prem K, Liu Y, Russell TW, Kucharski AJ, Eggo RM, Davies N, et al. The effect of control strategies to reduce social mixing on outcomes of the COVID-19 epidemic in Wuhan, China: a modelling study. Lancet Public Heal. 2020;5: e261–e270. doi:10.1016/S2468-2667(20)30073-6

22. Anderson RM, Heesterbeek H, Klinkenberg D, Hollingsworth TD. How will country-based mitigation measures influence the course of the COVID-19 epidemic? The Lancet. Lancet Publishing Group; 2020. pp. 931–934. doi:10.1016/S0140-6736(20)30567-5

23. Zimmermann P, Curtis N. Coronavirus infections in children including COVID-19: An overview of the epidemiology, clinical features, diagnosis, treatment and prevention options in children. Pediatric Infectious Disease Journal. Lippincott Williams and Wilkins; 2020. pp. 355–368. doi:10.1097/INF.0000000000002660

24. Gaunt ER, Hardie A, Claas ECJ, Simmonds P, Templeton KE. Epidemiology and Clinical Presentations of the Four Human Coronaviruses 229E, HKU1, NL63, and OC43 Detected over 3 Years Using a Novel Multiplex Real-Time PCR Method. J Clin Microbiol. 2010;48: 2940–2947. doi:10.1128/JCM.00636-10

25. Wang J, Tang K, Feng K, Lv W. High Temperature and High Humidity Reduce the Transmission of COVID-19. SSRN Electron J. 2020. doi:10.2139/ssrn.3551767

26. Ficetola GF, Rubolini D. Climate affects global patterns of COVID-19 early outbreak dynamics. medRxiv. 2020; 2020.03.23.20040501. doi:10.1101/2020.03.23.20040501

27. Bock Axelsen J, Yaari R, Grenfell BT, Stone L. Multiannual forecasting of seasonal influenza dynamics reveals climatic and evolutionary drivers. [cited 24 May 2020]. doi:10.1073/pnas.1321656111

28. HOUSEHOLDS AND FAMILIES: DEMOGRAPHIC CHARACTERISTICS 2018 Based on Labour Force Surveys. [cited 8 Jun 2020]. Available: https://www.cbs.gov.il/en/publications/Pages/2020/HOUSEHOLDS-FAMILIES-LabourForce-2018.aspx

29. (No Title). [cited 8 Jun 2020]. Available: https://www.idi.org.il/media/12168/the-yearbook-of-ultra-orthodox-society-in-israel-2018-he.pdf

30. Mossong J, Hens N, Jit M, Beutels P, Auranen K, Mikolajczyk R, et al. Social contacts and mixing patterns relevant to the spread of infectious diseases. PLoS Med. 2008. doi:10.1371/journal.pmed.0050074

31. Zhou F, Yu T, Du R, Fan G, Liu Y, Liu Z, et al. Clinical course and risk factors for mortality of adult inpatients with COVID-19 in Wuhan, China: a retrospective cohort study. Lancet. 2020;395: 1054–1062. doi:10.1016/S0140-6736(20)30566-3

32. Coronavirus (COVID-19) Data Dashboard - Novel Coronavirus (COVID-19) - County of Santa Clara. [cited 24 May 2020]. Available: https://www.sccgov.org/sites/covid19/Pages/dashboard.aspx

33. Italy Coronavirus: 232,664 Cases and 33,340 Deaths - Worldometer. [cited 30 May 2020].Available: https://www.worldometers.info/coronavirus/country/italy/

34. Li W, Zhang B, Lu J, Liu S, Chang Z, Cao P, et al. The characteristics of household transmission of COVID-19. [cited 24 May 2020]. doi:10.1093/cid/ciaa450/5821281

35. Dowell SF, Shang Ho M. Seasonality of infectious diseases and severe acute respiratory syndrome - What we don’t know can hurt us. Lancet Infectious Diseases. Lancet Publishing Group; 2004. pp. 704–708. doi:10.1016/S1473-3099(04)01177-6

36. Subjects - Live Births. [cited 8 Jun 2020]. Available: https://www.cbs.gov.il/en/subjects/Pages/Live-Births.aspx

37. COVID-19 Datasets-Government Data. [cited 30 May 2020]. Available: https://data.gov.il/dataset/covid-19

